# Quantifying within-school SARS-CoV-2 transmission and the impact of lateral flow testing in secondary schools in England

**DOI:** 10.1101/2021.07.09.21260271

**Authors:** Trystan Leng, Edward M. Hill, Alex Holmes, Emma Southall, Robin N. Thompson, Michael J. Tildesley, Matt J. Keeling, Louise Dyson

**Author notes:** https://maths.org/juniper/.

## Abstract

**Background:** To control within-school SARS-CoV-2 transmission in England, secondary school pupils have been encouraged to participate in twice weekly mass testing via lateral flow device tests (LFTs) from 8th March 2021, to complement an isolation of close contacts policy in place since 31st August 2020. Strategies involving the isolation of close contacts can lead to high levels of absences, negatively impacting pupils.

**Methods:** We fit a stochastic individual-based model of secondary schools to both community swab testing data and secondary school absences data. By simulating epidemics in secondary schools from 31st August 2020 until 21st May 2021, we quantify within-school transmission of SARS-CoV-2 in secondary schools in England, the impact of twice weekly mass testing on within-school transmission, and the potential impact of alternative strategies to the isolation of close contacts in reducing pupil absences.

**Findings:** The within-school reproduction number, *R*_*school*_, has remained below 1 from 31st August 2020 until 21st May 2021. Twice weekly mass testing using LFTs have helped to control within-school transmission in secondary schools in England. A strategy of serial contact testing alongside mass testing substantially reduces absences compared to strategies involving isolating close contacts, with only a marginal increase in within-school transmission.

**Interpretation:** Secondary school control strategies involving mass testing have the potential to control within-school transmission while substantially reducing absences compared to an isolation of close contacts policy.

## 1 Introduction

The ongoing Coronavirus-2019 (COVID-19) pandemic has seen unprecedented social restrictions placed upon populations globally. These have included general social distancing measures, the prohibition of households mixing socially, travel restrictions, the closure of pubs, restaurants, and non-essential shops, and have often involved the closure of schools. However, given the importance of school attendance in future academic attainment, employment prospects, and income^1,2^, school closures have been seen as a last resort ^3^, with schools remaining open throughout some periods of lockdown in England. Further, school closures may exacerbate educational inequalities ^4,5^, negatively impact children’s mental health ^6^, and reduce access to much needed services for the most vulnerable children ^7^. As countries emerge from a lockdown situation with the hope of relaxing social restrictions entirely, in the context of increasing immunity in the population through vaccine uptake ^8^, the question becomes how to minimise within-school transmission while keeping schools open, whether this can be done using strategies that minimise the disruption caused by isolating close contacts of individuals who test positive, and whether implemented school-level control measures have been effective at reducing transmission.

While COVID-19 infection rarely results in acute adverse health consequences for children^9–11^, preliminary data suggests a substantial proportion of cases in this age group exhibit symptoms consistent with long COVID for a period of months after infection ^12,13^. Moreover, controlling transmission within this age group remains important because of potential onwards transmission to families, teachers and the wider community. While previous studies tentatively suggest that transmission within schools are not driving transmission in the community ^14,15^, measures implemented at the school level may sometimes be sufficient to reduce the reproduction number (*R*) below 1, when there are no further community (non-school) based measures that can be implemented (or when further community based measures are undesirable) ^16^. Other studies have found that multiple within-school control measures in combination can be capable of mitigating within-school transmission ^17,18^ and the risk of onwards transmission from schools to the community caused by schools reopening ^19^.

In England, a range of school-level policies have been implemented to minimise transmission of the SARS-CoV-2 virus within schools ^20^. In secondary schools, measures applied have included mask-wearing for pupils and teachers (mandated from 8th March 2021 until 17th May 2021^21^), strict social distancing implemented through seating plans and the restriction of movement around schools, the implementation of ‘bubbling’ policies at the level of year groups or classes (to minimise the number of interaction and hence transmission opportunities), and the temporary isolation of infected individuals and close contacts upon confirmation of a positive case.

Alongside these measures, since the reopening of secondary schools in England on 8th March 2021, both teachers and secondary school pupils have been strongly encouraged to participate in twice weekly mass testing using lateral flow device tests (LFTs). By rapidly identifying asymptomatic and presymptomatic individuals, the aim of this strategy has been to minimise the increase in transmission associated with keeping schools open. Initially, participating pupils were tested in school three times; after this, tests were conducted at home. Any positive tests identified through home testing have been followed up by a confirmatory polymerase chain reaction (PCR) test, to minimise unnecessary absences from false positives. This policy has operated in tandem with a policy of isolation of close contacts of cases, to halt chains of transmission from infections that may have already taken place.

While PCR tests must be processed in a laboratory, meaning results typically take up to 48 hours to return, LFTs can be taken at home and are capable of returning a result within 30 minutes. The rapidness of LFTs make them ideal candidates for mass testing, and in the UK these have also been offered to the population at large since 9th April 2021. However, compared to PCR tests, LFTs are both less sensitive and less specific ^22,23^. Despite their comparatively lower sensitivity, there is evidence emerging that such tests can play an important role in rapid testing; previous studies have found that when employing a mass testing strategy, test sensitivity is secondary to testing frequency for reducing transmission ^24^. A more targeted use of LFTs within schools, known as serial contact testing, has also been suggested as a way to reduce transmission within schools that does not result in high levels of absences. Under serial contact testing, the close contacts of positively identified pupils are tested daily using LFTs for the next seven days, instead of isolating for ten days. Pilots within secondary schools in England to determine the efficacy of serial contact testing have been undertaken ^25^, with a serial contact testing policy considered as part of a national secondary school reopening strategy prior to the emergence of the Alpha (B.1.1.7) variant^26^.

Previous studies have attempted to capture the impact of school-based measures on within-school transmission. In previous work, we considered the impact that strategies involving LFTs could have on both transmission and absences in secondary schools ^27^, finding that serial contact testing alone would be insufficient to control within-school transmission, but a policy of regular mass testing alongside serial contact testing could be more effective than the isolation of year group bubbles while reducing absences considerably. A parallel study by Kunzmann *et al*. considered the impact of such measures in primary schools, reaching a similar conclusion that serial contact testing alone would be insufficient to contain outbreaks, recommending a combination of mass testing and isolation of close contacts ^28^. Other studies have considered the effectiveness of strategies involving PCR tests ^29^, the impact of mask-wearing on within-school transmission ^30^ and the benefits of dividing classes into discrete cohorts ^31^. Owing to the paucity of information surrounding the transmission rates of SARS-CoV-2 between children within schools, such studies typically consider a range of within-school transmission rates. While this approach is reasonable and valuable to compare the relative impact of different school-based measures, quantifying the impact of implemented control measures on transmission requires realistic levels of within-school transmission. Other studies have considered the impact of school closures or school control strategies on transmission in the wider community ^16,32–34^, often by altering the age-dependent mixing matrices assumed in the underlying model. While these studies are important in understanding the impact that schools have on community transmission, such methods are insufficient to quantify the impact of control measures on within-school transmission.

In this study, we extend our previously described stochastic individual-based model of a secondary school formed of exclusive year-group bubbles ^27^. Specifically, we incorporate realistic secondary school sizes and close contact group sizes, derived from Department for Education: Educational Setting Status data ^35^, we use S-gene negative data to incorporate the spread of the Alpha (B.1.1.7) variant and its impact on within-school transmission, and we use swab testing data from the wider population ^36^, referred to as Pillar 2 data, to inform each school’s probability of external infection from the local community and the uptake of LFTs, based on each school’s lower tier local authority (LTLA). We fit this model using an Approximate Bayesian Computation (ABC) approach ^37^ to positive PCR and LFT time-series data of 11-16 year olds and the distribution of peak confirmed COVID-19 cases in secondary schools. From this fitted model, we estimate the proportion of infections in secondary school pupils that occur due to within school transmission, the school reproduction number (*R*_*school*_), the impact of LFTs on incidence at current levels of uptake, the benefit of higher levels of LFT uptake, and the potential impact of serial contact testing instead of isolating close contacts. These analyses highlight approaches in which transmission within the school environment can be kept low while maintaining high levels of attendance, a difficult balance that is vitally important if we are to preserve the benefits of education during future waves of the pandemic.

## 2 Methods

We extend our stochastic individual-based model of secondary schools formed of year-group bubbles, detailed in full previously ^27^, to first quantify the level of within-school transmission of SARS-CoV-2 in secondary schools in England from 31st August 2020 to 21st May 2021, and then assess the impact of LFTs on within-school transmission from 8th March 2021 to 21st May 2021.

We describe the data sources used within this study in Section 2.1, overview the individual-based model in Section 2.2 and outline the model fitting procedure in Section 2.3. In Section 2.4, we consider alternative school reopening strategies (other than isolation of contacts) and, in Section 2.5, give our outcome measures used to asses the impact of different strategies on within-school transmission and absences. We performed the model fitting, model simulations, and visualisation of results using MATLAB 2019b.

### 2.1 Data summary

#### Community swab testing

Since the beginning of the COVID-19 outbreak, the UK has collected and recorded daily testing data ^36^. Pillar 2 data refers to community swab testing data from those who have sought PCR tests due to COVID-19 symptoms, or from those undertaking LFT tests. We fit our model to two strands of Pillar 2 data: (i) the proportion of 11-16 year olds testing positive to a PCR test each week (excluding confirmatory PCR tests, which are accounted for in LFT testing data) throughout both terms, and (ii) the proportion of 11-16 year olds testing positive to an LFT test each day from the 8th March 2021. We focus on the 11-16 year olds age group as the vast majority of 11-16 year olds attend secondary schools in England; we therefore view that these data can be used as a proxy for testing rates within secondary school pupils. We excluded 17-18 year olds as some of these will be employed or attending university, and therefore trends may emerge in testing data in these age groups that do not reflect the dynamics within secondary schools, but rather reflect dynamics in a different setting (e.g. a university). These data also informed the level of LFT uptake in each LTLA in the model. The uptake in each LTLA on a given day was taken as the proportion of 10-19 year olds in that LTLA who recorded either a positive or negative LFT on that day. We used the broader age range of 10-19 year olds here due to the negative Pillar 2 data only being available in five year age bands. It is likely that a considerable proportion of negative home tests remain unrecorded; to account for this we fit the level of underreporting of negative tests, which scales up the ‘true’ number of negative LFT tests and hence increases uptake.

The Alpha (B.1.1.7) variant is characterised by a deletion in the genome at site 69-70 associated with the spike protein; this lead to the ThermoFisher TaqPath quantitative PCR assay, used as the main diagnostic tool in many regions of England, failing to amplify the S-gene target. As such infections that are confirmed by PRC but where the S-gene is not detected (often termed as ‘S-gene failures’) provide a key indicator for the geographic spread of the Alpha variant. We therefore use the change in proportion of S-gene failures within a LTLA as a proxy for the sigmoidal growth of the Alpha variant which was associated with higher transmission rates than the original wild-type variant.

#### Population by LTLA

We obtained the population sizes in each LTLA from the Office for National Statistics population estimates, using the mid-year estimates from 2019^38^. We assumed that 11-16 year olds who tested positive in a particular LTLA also attended school in that LTLA.

#### School absence data

Since September 2020, schools have recorded data regarding absences and confirmed COVID-19 cases, available through the Department for Education: Educational Setting Status data ^35^. In particular, each school has recorded the number of absences due to confirmed COVID-19 cases among secondary school pupils each day. While these data are insufficient to establish the total number of cases there have been in any particular school, one can obtain the peak number of confirmed cases in each school. To capture the heterogeneity in outbreak sizes between schools, we fitted our model to the distributions of peak number of confirmed cases in schools from September to December 2020 and from March to May 2021. Secondary schools were closed to the majority of pupils in January and February 2021, as part of a national lockdown, and because of these closures we did not consider confirmed cases in schools in this period.

In addition to the number of absences due to a confirmed case of COVID-19, also recorded were the number of absences due to other within-school COVID-19 related reasons. We define such an absence as an absence due to a confirmed case of COVID-19, an absence due to a suspected case of COVID-19, or an absence due to potential contact with a case of COVID-19 from inside the educational setting (and so the student was requested to isolate). Prior to 10th October 2020, the number of pupils that were asked to isolate due to potential contact with a case of COVID-19 were not separated into contact with a case from within the educational setting, and contact with a case from outside the educational setting. As such, we are overestimating the total number of absences for within-school COVID-19 related reasons over this period.

### 2.2 Model description

#### School contact structure

Our previous study described a situation where secondary schools implemented a bubbling policy at the level of year groups, and assumed that within year group pupils mixed randomly, to understand the impact of control measures targeted at year groups ^27^. However, while the majority of secondary schools report implementing a bubbling policy at the level of year groups ^39^, schools have often isolated smaller groups of targeted close contacts upon confirmation of a positive case ^40^, in line with the guidance to secondary schools in England as of June 2021^20^. To account for this, we extended our model to consider three levels of mixing. Each year group was comprised of exclusive sets of close contacts of equal size, with each school comprised of several year groups. We set pupils to mix with their close contacts at a rate *α*_0_, mix with pupils in the rest of their year group at a rate *α*_1_, and mix with pupils in the rest of their school at a rate *α*_2_ (Figure 1a). By default, we set *α*_0_ = 1 and let 0 *≤α*_2_, *α*_1_ *≤*1. The hierarchy of *α* terms reflects relative mixing rates compared to a baseline mixing rate of close contacts. Letting *I*_*i*_(*d*) denote the infectiousness of individual *i* on day *d*, and letting *N*_*x*_ denote the number of contacts a pupil has of type *x*, the probability of within-school transmission to a susceptible individual *j, β*_*j*_(*d*) is given by:

**Figure 1:**
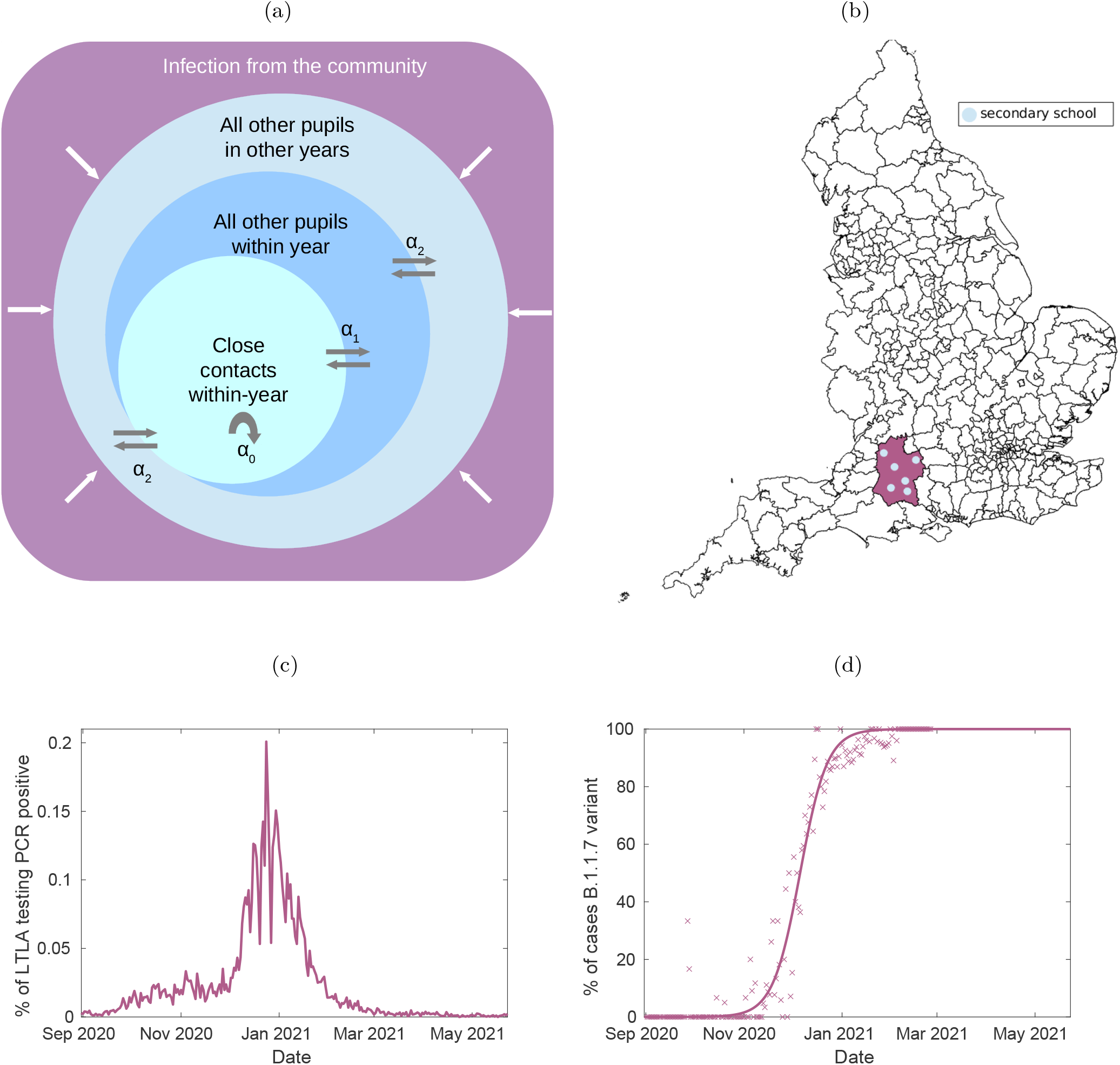
Overview of the individual-based model components. (a) A schematic of the within-school mixing structure assumed in the model. Within a school, pupils interact with close contacts in their year at a rate *α*_0_ = 1, with other pupils within their year at a relative rate *α*_1_, and interact with other pupils in other years at a relative rate *α*_2_, where 0 *≤α*_2_, *α*_1_ *≤* 1. (b) A map of England segregated by LTLA, with an example LTLA highlighted in purple. Each school is situated within an LTLA, which determines its probability of infection from the community, its relative frequency of B.1.1.7 (Alpha) variant, and LFT uptake. Each LTLA contains multiple secondary schools, shown as blue dots (the number of blue dots shown is illustrative rather than an accurate depiction of the number of secondary schools in the highlighted LTLA). (c) A time-series of the percentage of that LTLA’s population who test positive to a PCR test on that day. A pupil’s probability of external infection on day *t* depends upon prevalence in the community, which we assume to be proportional to the proportion of the population in that LTLA testing PCR positive on day *t* + 5. (d) A time-series of the fitted estimate of the relative frequency of the B.1.1.7 variant in the example LTLA. The expected number of secondary infections from infected pupils depends upon the proportion of cases that are of the (more transmissible) B.1.1.7 variant, which varies through time and is dependent on the LTLA the school is situated within. Cross markers indicate the percentage of sequenced tests from an LTLA that return an S-gene negative result. Our model does not consider the impact of the B.1.617.2 (Delta) variant, which became the dominant variant in circulation during late May 2021, occurring beyond the time horizon considered in our analyses.

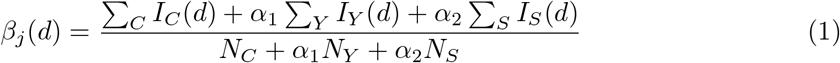

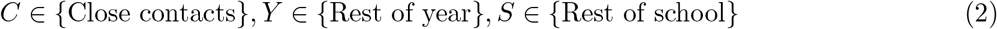

such that *α*_1_ = *α*_2_ = 1 corresponds to random mixing across the entire school, while at the other extreme, where *α*_1_ = *α*_2_ = 0, pupils mix exclusively with their close contacts. In our baseline scenario, we assumed *α*_1_ = 0.1 and *α*_2_ = 0.01, reflecting a situation where pupils have a much higher rate of interaction with their close contacts than other members of their year group, and where year group bubbles have been in general very effective (with only a small rate of interaction to account for the impact of indirect infection via teachers and siblings). To understand the sensitivity of our results to these mixing assumptions, in Supporting Text S7, we consider a situation where mixing within-years and across years occurs to a much higher extent, setting *α*_1_ = 1 and *α*_2_ = 0.1, with qualitatively similar results obtained.

#### Transmission

This section provides an overview of the transmission dynamics assumed in the model, with an extended description of the model’s transmission dynamics described in Supporting Text S2.

Within the model, an infected pupil’s relative probability of transmission to other pupils since day of infection was based upon a previously derived infectivity profile for COVID-19^41^, based on data from known source-recipient pairs ^42^, with an assumed incubation period distribution under the assumption that the generation time and incubation period are independent. Symptomatic pupils developed symptoms on a day drawn from a Gamma distribution with shape 5.807 and scale 0.948^43^, corresponding to a mean time to symptom onset of 5.5 days. Many secondary school aged children remain asymptomatic over the course of their infection ^44^. Further, asymptomatic individuals are likely less infectious than symptomatic individuals ^45,46^, and so asymptomatic transmission is included explicitly in the model.

The initial prevalence of each school at the start of the simulation depended on that school’s LTLA, while the initial level of immunity at the start of simulation was set as 6.25%, reflecting the estimated prevalence of COVID-19 antibodies in September 2020 nationally ^47^.

Within the model, the rate of external infection of pupils depended on each school’s LTLA and varied through time. Detailed temporal COVID-19 testing data has been recorded at the LTLA level ^36^, and the positive testing rate within a population is a function of the prevalence in that population. To account for the delay between contracting infection and displaying symptoms, with a mean time to symptom onset of 5.5 days, we assumed that the rate of external infection of school pupils on day *t* was proportional to the proportion of all individuals in that school’s LTLA testing positive to a PCR test on day *t* + 5. This positive testing rate was then scaled by a factor depending on whether a school was situated in a rural or urban LTLA, reflecting that children in rural communities likely have less exposure to community infection than pupils in urban areas. For all schools, the probability of external infection was scaled by a factor *h* during school holidays.

While the impact of more transmissible variants on external transmission is captured through the above set-up, such variants also impact the level of onward transmission from infected pupils within schools. Over the course of schools reopening, the B.1.1.7 variant emerged and subsequently dominated infections across England ^48^. The B.1.1.7 variant returns a negative result for the spike protein gene when tested using the Thermo TaqPath PCR system (referred to as S-gene negative), while other variant in circulation at the time do not (S-gene positive). To capture the impact of this variant on within-school transmission, we therefore scaled the within-school transmission parameter *K*_*s*_ according to the proportion of sequenced tests in each LTLA that return an S-gene negative result through time (Supporting Figure S2). The increase in transmission from this parameter was determined through the fitting procedure for the model. Our analysis does not consider the impact of the B.1.617.2 variant, commonly referred to as the Delta variant, which began to dominate new infections across England from May 2021^49^.

In total, the model fits six parameters relevant to within-school transmission, and four parameters relevant to community infection (Supporting Table S1). In a given secondary school *s*, within-school transmission was determined by a parameter *K*_*s*_, drawn from a lognormal distribution, the parameters of which are determined during fitting. Alongside these, we fit the increase in within-school transmission resulting from increased prevalence of the B.1.1.7 variant, and the increase in within-school transmission after the 26th October-30th October 2020 half term, to capture the impact of reduced adherence to strict within-school distancing measures implemented at the beginning of term. We determined via the fitting procedure both the proportion of infected pupils that remain asymptomatic and the relative infectiousness of asymptomatic individuals. We also fit a scaling factor between community testing rates and the probability of external infection in school *s, E*_*S*_, drawn from a lognormal distribution, the parameters of which are determined during fitting. For rural LTLAs, this was scaled further by a factor *r*, while another scaling factor *h* impacted pupils’ probability of infection during periods of school holidays/closures, with values for *h* and *r* determined during fitting.

#### Testing and control

Throughout the simulation, infected pupils underwent a PCR test upon symptom onset. Pupils self-isolated until they received a test result. We assumed that pupils received a result two days after taking a test (though in reality there will be some variation in the length of time between taking a test and receiving a result). Those receiving a negative result returned to school the day after receiving this notification, while those testing positive entered isolation for a period including the day that symptom onset began and the next ten full days ^50^. Pupils who tested positive using an LFT entered isolation, with the outcome of a confirmatory PCR test then determining whether the pupil remained in isolation (for a period including the day the LFT test was taken and the next ten full days ^50^). Twice weekly mass testing in schools began on the 1st March 2021, i.e. the week before schools reopened. In each school, each pupil was assigned two days of the week (Sunday and Wednesday, Monday and Thursday, etc.) in which they were scheduled to undertake an LFT test each week. The probability of those pupils taking a test on that given day was matched to satisfy the uptake of LFTs in that school’s LTLA (Supporting Text S3). As LFTs have predominantly been taken at home, we assumed that identified infected pupils did not transmit infection on the day they undertook a test. We used previously estimated LFT and PCR test probability profiles for symptomatic individuals ^51^. For asymptomatic individuals, we assumed that the probability of testing positive was equal to that of symptomatic individuals until peak positive test probability, but then decayed more rapidly, as in our previous study ^27^. Both the specificity of LFTs and the proportion of negative home tests taken that go unreported were determined through the fitting procedure. The specificity of PCR tests was assumed to be 1, in line with data indicating that false PCR positives are very rare ^52^.

Secondary schools isolated the close contacts of infected pupils upon a pupil testing positive to a PCR test (either through self-seeking or as a confirmatory test to a positive LFT) for ten days following the day of last contact ^53^. Within the model, the size of close contact groups for each school remained constant over each term, informed by reported absences data. To reflect that schools have in general isolated smaller groups of pupils from March 2021 onwards, the size of close contact groups therefore differed between September to December 2020 and March to May 2021 (Supporting Figure S1b). We describe the derivation of modelled close contact sizes and modelled secondary school sizes in Supporting Text S1.

### 2.3 Model fitting

The model is fitted using an Adaptive Population Monte Carlo ABC approach ^37^. Initially, we sampled 100 particles from the prior distribution (Supporting Table S1. In each generation, we retained the top 20% of particles, which informed the covariance matrix for the subsequent generation. Initially, we performed the fitting for a sample of 100 schools for ten generations, to navigate parameter space efficiently. We then ran the model fitting scheme for a sample of 1000 schools until we observed no improvement in the median log likelihood obtained per generation, a total of 21 generations (Supporting Figure S5).

We fitted the model to Pillar 2 testing data and peak confirmed cases in schools data outlined above. The log likelihood of the model given the data could be defined for each of the data streams. We based the time-series components of the log-likelihood function on a binomial likelihood for each day. We let *X*_*T*_ (*d*) denote the observed number of positive tests of type *T, T ∈* {PCR, LFT} in 11-16 year olds on day *d, Y*_*T*_ (*d*) denote the predicted proportion of pupils who test positive on day *d* to a test of type *T*, and we let *N* denote the population size of 11-16 year olds in England. *L*_*B*_(*n*|*N, p*) denote the log of the binomial probability function. We assumed the components corresponding to the distributions of peak cases in schools followed a multinomial distribution. We let *S*_*P*_ (*k*) denote the number of secondary schools with a peak of *k* confirmed cases over period *P, P∈* {September to December 2020 (*septodec*), March to May 2021 (*martomay*)}, *Y*_*P*_ (*k*) denote the predicted proportion of schools with a peak of *k* cases over period *P*. The log-likelihood function obeyed:

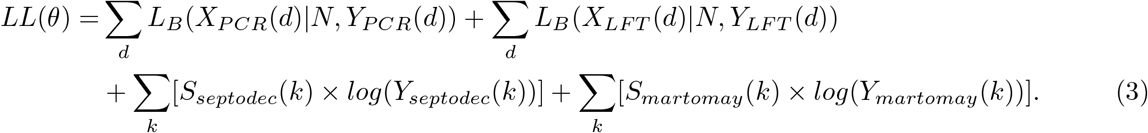

### 2.4 Alternative strategies

Using the parameters obtained from our model fitted, fit to the observed data during which the interventions measures used were isolating close contacts and twice weekly mass testing, we assessed the impact that other potential strategies would likely have had on within-school transmission and absences (had the alternative package of interventions been introduced for schools fully reopening on 8th March 2021) through model simulation. We focused on three such alternative strategies:

1. *Isolation of close contacts only*. A counterfactual scenario, with no use of mass testing. Schools continue to implement the isolation of close contacts policy used by schools from 31st August-18th December 2020.
2. *Mass testing only*. From the 1st March 2021, the week prior to schools fully reopening, pupils undertake twice weekly mass testing (calibrated to obtain realistic levels of uptake). However, identification of a positive case led to no further action, other than isolating the confirmed positive individual.
3. *Mass testing and serial contact testing*. Alongside mass testing, and upon identification of a positive case via a PCR test (either from a symptomatic pupil seeking a PCR test or as a confirmatory test from a positive LFT), that pupil’s close contacts take LFTs for the next seven days following the last contact with the positive case (serial contact testing). It is assumed that all pupils participate in serial contact testing upon being identified as a close contact of a positive case.

### 2.5 Outcome measures

To understand the impact of different strategies on within-school transmission and pupil absences, we used three main outcome measures:

- *Incidence*. Tracking the incidence, and the impact of control measures on incidence, is a natural measure to judge the benefit of control measures in reducing infections. However, incidence also depends on within-school prevalence and community prevalence, so does not (directly) inform us whether transmission is under control in secondary schools. Further, by tracking whether new infections occur through a within-school contact or through external contact, we can estimate the proportion of infections that occur within-school during term time, and whether this has changed over the course of schools reopening.
- *Within-school reproduction number (R*_*school*_). A case reproduction number, defined as the number of secondary infections resulting from contact with an individual infected on day *t* divided by the number of individuals infected on date *t*. This outcome measure tells us whether within-school transmission is under control, indicated by *R*_*school*_ *<* 1, and how this has changed through time in the context of emerging variants and changing control measures.
- *Percentage of pupils absent*. The percentage of all modelled pupils absent on a given day, either because they have tested positive or because they are a close contact of a positively identified individual in a school implemented an isolation of close contacts policy. This outcome measure is useful to understand the impact that different strategies have regarding pupil absences and the potential disruption to pupil attendance.

## 3 Results

### 3.1 Model fit and parameter inference

The fitted model matches well to the temporal data on PCR and LFT positivity (Figures 2a and 2b), whilst also providing a reasonable fit to the distribution of peak case numbers across schools in the September-December 2020 and March-May 2021 periods (Figures 2c and 2d). We acknowledge an underestimation in the model-fitted proportion of schools that had a low peak number of confirmed cases during September-December 2020 (Figure 2c), while an overestimation in the proportion of schools with a low peak number of confirmed cases during March-May 2021 (Figure 2d).

**Figure 2:**
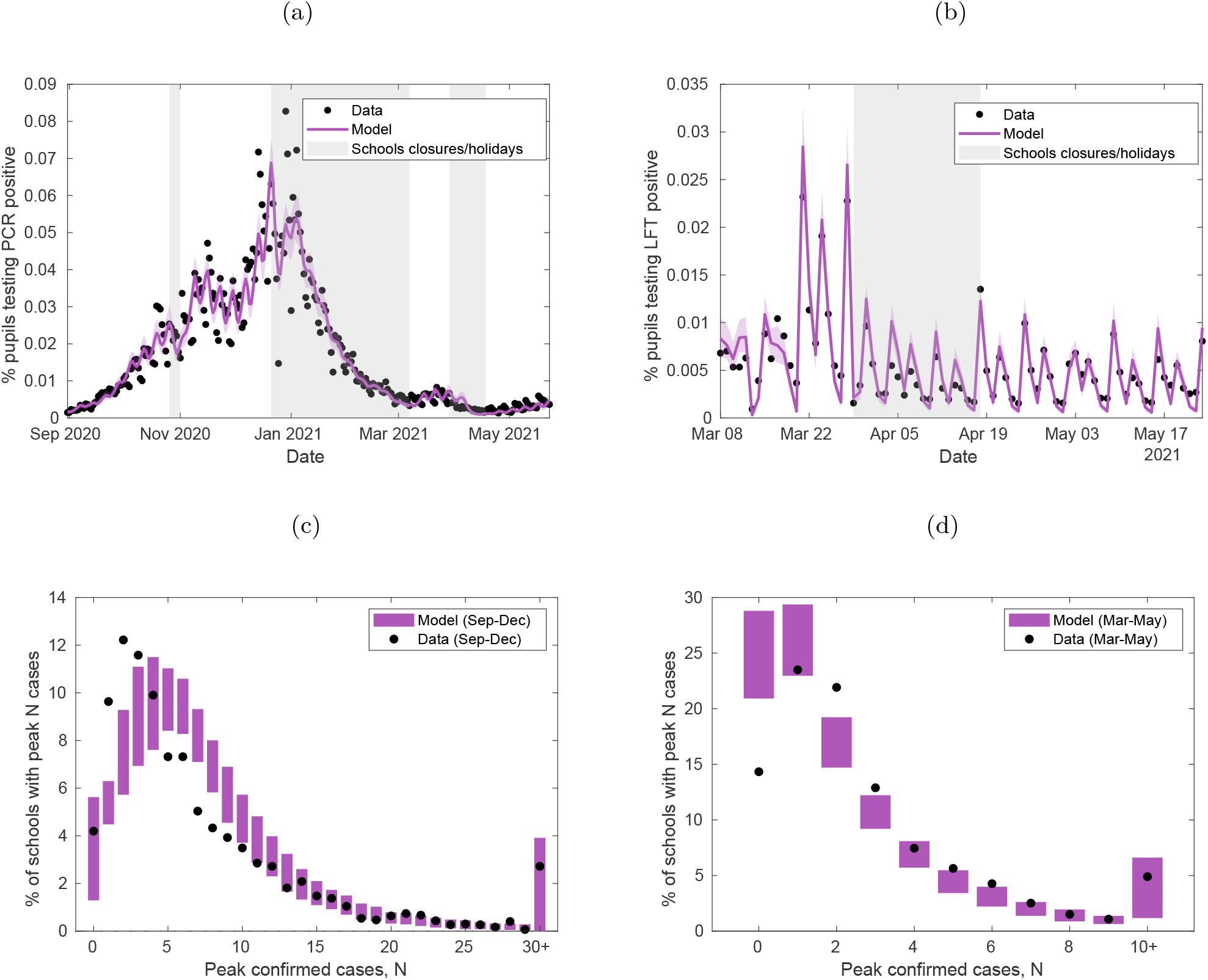
Fitting the model to testing and secondary school absences data. The stochastic individual-based model is fitted to (a) the percentage of 11-16 year olds who test PCR positive (excluding confirmatory PCR tests) each day from 1st September 2020 to 21st May 2021, (b) the percentage of 11-16 year olds who test LFT positive each day from 8th March 2021 to 21st May 2021. Shaded intervals around mean model traces (solid lines) represent 95% prediction intervals in all plots, while shaded grey regions represent time periods when schools were not fully open (either due to closures or school holidays). The model is also fitted to (c) the distribution of peak number of confirmed COVID-19 absences in secondary schools from 1st September 2020 to 18th December 2020, and (d) the distribution of peak number of confirmed COVID-19 absences in secondary schools from 8th March 2021 to 21st May 2021. Filled circles denote the data and shaded blocks the 95% prediction interval estimated from the model. The plots above show the mean values obtained from 100 simulations of 2979 secondary schools, each with a distinct parameter set sample from the posterior distribution.

We infer an increase in within-school transmission to the B.1.1.7 variant in secondary-school aged children of approximately a mean value of 72% (95% credible interval: 58-84%), and attribute a 30% (95% credible interval: 13-45%) increase in within-school transmission after schools return in the second half-term of the October-December term, due to falling adherence to within-school control measures over time. The data are best explained by a model that assumed only 39% (95% credible interval: 31-54%) of negative home LFTs are in fact reported. Pupils attending schools in rural LTLAs had a considerably lower probability of external infection than those attending schools in urban LTLAs, with community testing rates in rural schools scaled by a factor 34% (95% credible interval: 11-55%) of the scaling factor for urban schools to obtain a school’s probability of external infection. The model results in similar levels of COVID-19 related absences to the observed level of absences in Department for Education: Education Setting Status data (Supporting Figure S6). As well as matching testing data nationally, the model also matches PCR testing data closely at a regional level (Supporting Figure S7), although underestimates the heterogeneity in PCR and LFT testing data between LTLAs (Supporting Figure S8). A model fitted under alternative within-school mixing assumptions was also capable of matching both streams of data reasonably well (Figure S9).

### 3.2 Quantifying within-school transmission

For our analysis of the proportion of transmission events attributed to within-school contacts rather than external settings, we found the majority of pupil infections during term time on school days occurred within school, as opposed to extraneously from the community (Figure 3a). The proportion of cases occurring within school increased through time, accounting for 49% (95% credible interval: 41-58%) of all new infections in the September-October half term, 72% (95% credible interval: 67-76%) of all new infections in the November-December half term, and 76% (95% credible interval: 73-79%) of infections since schools reopened from 8th March 2021 until 21st May 2021. This increasing trend in transmission within the school setting is echoed when considering the within-school reproduction number - while *R*_*school*_ remained well below 1 throughout the school term in 2020, its value rose to just below a value of 1 by May 2021, owing to the dominance of the B.1.1.7 strain in circulation (Figure 3b). Qualitatively similar results were obtained from a fitted model under the alternative within-school mixing assumptions *α*_1_ = 1, *α*_2_ = 0.1 (Supporting Figure S10).

**Figure 3:**
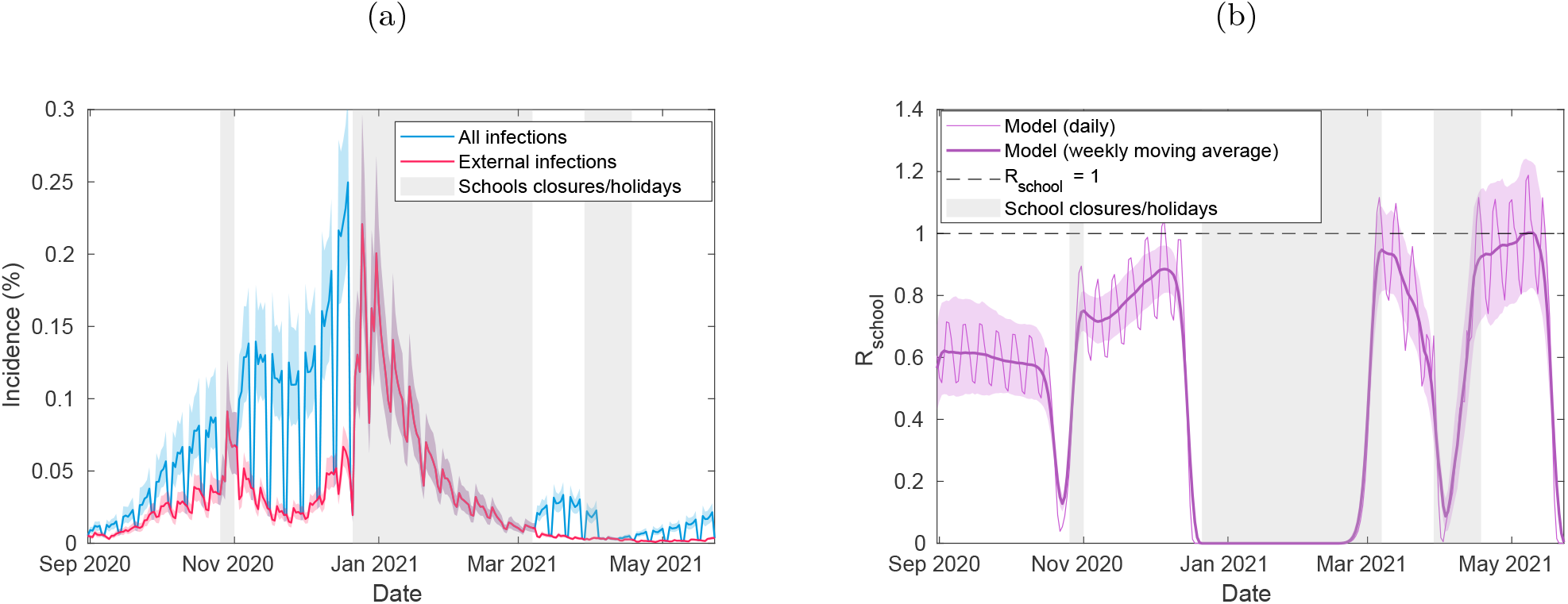
Incidence and *R*_*school*_ from the fitted model. We display time-series of (a) incidence among pupils, delineated into whether infections occur externally or within-school, and (b) *R*_*school*_ through time (thin line) alongside its seven-day moving average (thick line). The plots above were obtained from 100 simulations of 2979 secondary schools, each with a distinct parameter set sample from the posterior distribution. In all panels, solid lines correspond to mean temporal profiles, shaded ribbons represent 95% prediction intervals in all plots, while shaded grey regions represent time periods when schools were not fully reopen (either due to closures or school holidays).

### 3.3 Impact of LFTs

We compared our fitted model to a scenario where mass testing was not introduced upon schools reopening in March 2021. After accounting for underreporting of negative LFTs, there was an estimated 36% (95% credible interval: 28-42%) uptake nationally from 8th March until the 23rd May 2021, though LFT uptake varied substantially between LTLAs (Supporting Figure S3b). Despite the relatively low levels of LFT uptake, mass testing has reduced incidence within schools considerably (Figure 4a, purple line), compared to a scenario where mass testing has not been introduced (Figure 4a). However, at current levels of LFT uptake, mass testing is only just sufficient to keep *R*_*school*_ below 1 (Figure 4b, purple line).

**Figure 4:**
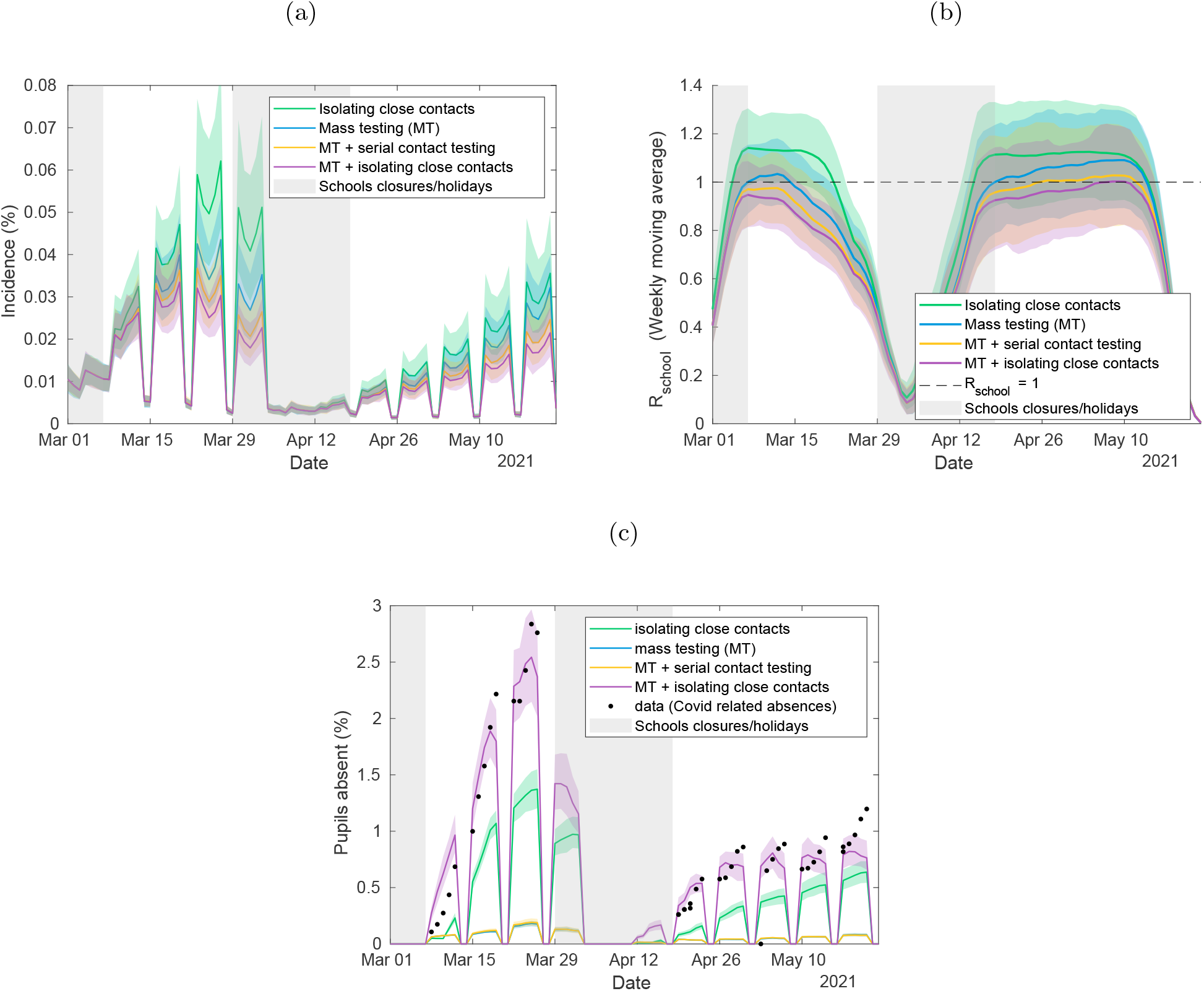
Quantifying the impact of LFTs on transmission and absences, and the potential impact of alternative strategies. Time-series under different intervention strategies of (a) incidence among pupils, (b) *R*_*school*_ within secondary schools, and (c) the percentage of pupils absent. We compare a policy of twice weekly mass testing and isolating close contacts (purple) to a strategy of isolating close contacts only (green), twice weekly mass testing only (blue), and twice weekly mass testing alongside serial contact testing (yellow). The plots above show the mean values obtained from 100 simulations of 2979 secondary schools, each with a distinct parameter set sample from the posterior distribution. In all panels, solid lines correspond to the mean estimate, shaded intervals represent 95% prediction intervals, while shaded grey regions represent time periods when schools were not fully reopen (either due to closures or school holidays). The data in Figure 4c consists of the number of absences due to a confirmed case or a suspected case of COVID-19, and absences arising as a result of students told to isolate due to potential contact with a case of COVID-19 from inside their educational setting. These data were recorded by 2979 secondary schools.

We can also compare our fitted model to a scenario where mass testing is introduced without close contacts being isolated, and to a scenario where serial contact tracing is operating instead of the isolation of close contacts. We found that, at current levels of uptake, mass testing alone would be insufficient to reduce *R*_*school*_ below 1(Figure 4b, blue line). Compared to the current strategy of isolating close contacts alongside mass testing, a strategy of serial contact testing alongside mass testing (Figure 4b, yellow line) was marginally less effective at controlling within-school transmission, indicated by the slightly higher values of *R*_*school*_ over the period considered. These strategies resulted in lower levels of within-school transmission than a strategy of isolating close contacts alone (Figures 4a and 4b).

Both such strategies were capable of reducing absences considerably when compared to either strategy involving isolation of close contacts (Figure 4c). Modelled COVID-19 related absences under the current strategy reached a peak of 2.54% (95% credible interval: 2.15-2.97%) over the period 8th March 2021 until 21st May 2021, in line with peak COVID-19 related absences observed in data. In contrast, a strategy of mass testing alone resulted in a peak of 0.18% (95% credible interval: 0.15-0.20%) of absences over the same period, while a strategy of mass testing alongside serial contact testing resulted in a peak of 0.19% (95% credible interval: 0.16-0.23%). Again, qualitatively similar results were obtained from a fitted model under the alternative within-school mixing assumptions *α*_1_ = 1, *α*_2_ = 0.1 (Supporting Figure S11).

### 3.4 LFT uptake counterfactuals

Finally, we considered whether higher levels of LFT uptake would be sufficient to bring *R*_*school*_ below 1. To understand the impact of uptake of twice weekly mass testing, we assumed that all pupils scheduled to take a test on a given day do so with a probability *p*, and we varied *p* from 0 to 1. Unsurprisingly, as uptake increases, *R*_*school*_ falls. Under a strategy of mass testing alongside isolation of close contacts, LFT uptake of 24% (95% credible interval: 8-39%) would have been required to bring the mean value of *R*_*school*_ below 1 from 19th April 2021 to 9th May 2021 (Figure 5).

**Figure 5:**
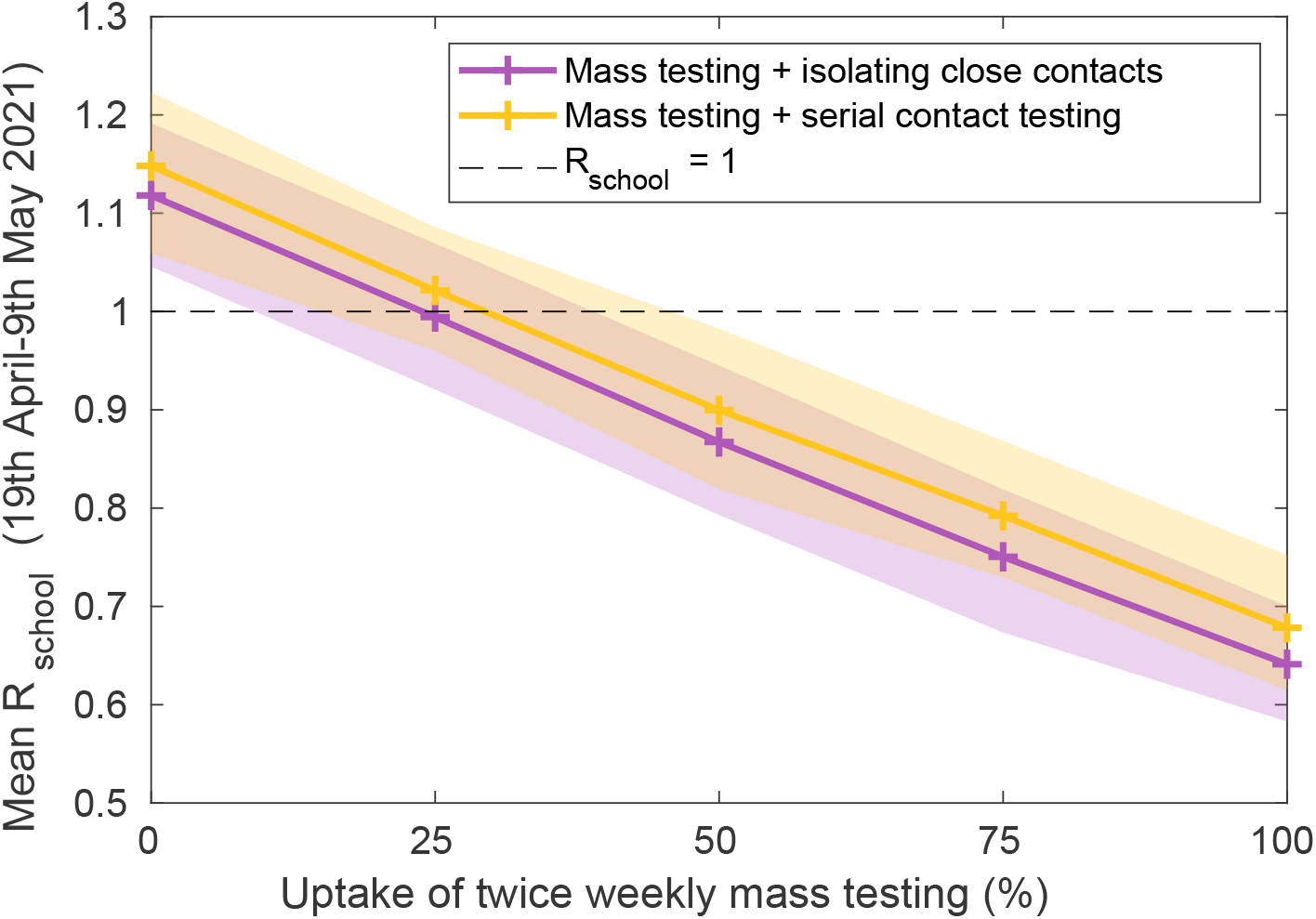
The impact of regular mass testing uptake on within-school transmission. We considered the impact of increasing uptake of twice weekly mass testing on the mean daily *R*_*school*_ from 19th April 2021 to 9th May 2021, for a strategy of twice weekly mass testing alongside the isolation of close contacts (purple), and for a strategy of twice weekly mass testing alongside serial contact testing (yellow). The plot above shows the mean values obtained from 100 simulations of 2979 secondary schools, each with a distinct parameter set sample from the posterior distribution, at uptakes of 0, 25, 50, 75, and 100% (indicated by the cross markers). Shaded intervals represent 95% prediction intervals.

We compared the impact of uptake for a strategy of isolation of close contacts against a strategy implementing serial contact testing alongside twice weekly mass testing. We assumed uptake of serial contact testing to be distinct from that of twice weekly mass testing, and assumed that 100% of identified close contacts participated in serial contact testing. Under the strategy including serial contact testing, keeping *R*_*school*_ below 1 required a slightly higher uptake of twice weekly mass testing 29%(95% credible interval: 14-45%). Both strategies considered were capable of reducing *R*_*school*_ below 1, predicated on a high uptake of twice weekly mass testing.

## 4 Discussion

Epidemiological models matched to available data are vital tools in understanding the impact of control measures. This study combines swab testing data collected from the wider population with absences data from secondary schools in England recorded from August 2020 to May 2021 to obtain a model of within-school transmission consistent with both streams of data. Doing so not only elucidates the impact of control measures that have been implemented – a combination of mass testing and isolation of school ‘bubble’ contacts – but can also be used to understand the potential impact of alternative strategies. Further, our model provides a quantification of within-school transmission, which can be used to inform future modelling work of realistic parameter values surrounding within-school transmission dynamics. Our research contributes to the growing epidemiological modelling literature regarding the role of schools in SARS-CoV-2 transmission in the UK^30,32,33,54,55^.

Our results demonstrate that a large proportion of cases in secondary-school aged children likely result from transmission within secondary schools, with such infections comprising approximately 49% of new infections in the September-October 2020 half-term, 72% of new infections in the November-December 2020 half-term, and 76% of new infections from 8th March-21st May 2021 in secondary-school aged children. At the same time, our results suggest that transmission was not ‘out of control’ within secondary schools over the course of the September to December 2020 term and from March to May 2021, as the estimated *R*_*school*_ was below Taken together, these results imply that sustained transmission within secondary schools has, over the time-period considered, required external infection from the community, a result consistent with previous research indicating that within-school transmission has not driven community infection ^14,15^.

While there appears to have been relatively low levels of LFT uptake in secondary schools in England (approximately 36%), this study demonstrates that LFTs have played an important role in reducing incidence within secondary schools, which will have consequently reduced incidence in the wider community. Our results underline the importance of such mass testing in controlling transmission within secondary schools, but also highlights the potential benefits of even higher levels of uptake. Our results therefore agree with an emerging picture that mass testing via LFTs is capable of playing an important role in reducing transmission ^24,27^, despite their lower sensitivity compared to PCR tests ^22^.

Our results reiterate our previous finding ^27^ highlighting that strategies involving the isolation of large numbers of close contacts lead to considerable levels of school absences. In the context of minimising disruption from pupil absences, we considered the likely impact of alternative strategies not involving isolation of close contacts. While mass testing alone (with an approximate uptake of 36%) would have been insufficient to keep *R*_*school*_ below 1 over the course of schools reopening from March to May 2021, a policy of regular mass testing alongside serial contact testing was almost as effective at reducing within-school transmission as regular mass testing alongside isolating close contacts, but considerably reduced absences.

When considering serial contact testing, this study assumed that all pupils agree to participate in daily testing upon being identified as a close contact of a positive case. In practice, some proportion of pupils are likely to not participate. If non-participating pupils who are identified as close contacts to infected individuals must self-isolate instead of taking daily tests, then lower uptake would likely not increase within-school transmission, but would increase pupil absences. Instead, if non-participating pupils are allowed to remain in school, absences would remain low but within-school transmission would increase. Which option is most suitable depends both on expected levels of participation and the intended goals of such a strategy, demonstrating that clear protocols and aims of strategies involving mass testing are paramount to their successful implementation ^56^.

Any mathematical modelling study is a simplification of the real-world, and necessarily makes a range of assumptions. Accordingly, our study has several limitations. Regarding transmission, our model captures the impact of community prevalence on within-school transmission, but does not capture the impact of within-school transmission on community prevalence. In reality, within-school epidemics may increase community prevalence in extremely local areas (smaller than that of an LTLA), which would then be expected to increase transmission in schools as a damped feed-back loop. Our study assumes homogeneous onwards transmission rates from infected pupils within a given secondary school. In reality, onwards transmission rates are likely heterogeneous between pupils, with transmission rates likely a function of viral load ^57^ which is heterogeneous both in time and between individuals. Previous studies accounting for this heterogeneity, through the incorporation of within-host viral dynamics ^28^ obtained similar results to our previous study ^27^, though the inclusion of heterogeneity may impact the cluster sizes of epidemics in schools ^58^.

Regarding testing, our model assumes that the proportion of pupils taking an LFT test on a given day is equivalent to the local proportion of 10-19 year olds in that school’s LTLA taking an LFT test on a given day, i.e. we assume uptake is homogeneous across schools within a region. In reality, there may be significant heterogeneity between schools even within a local area. Further, we assume that pupils have a given probability of taking an LFT test to satisfy a given level of uptake; in reality, there may be pupils who are consistently taking LFT tests, while there may be consistent non-adherence in others. Including such heterogeneities would be expected to increase the heterogeneity in cases between schools, with schools with lower testing uptake expected to have higher levels of within-school transmission. As a further complication, underreporting of negative tests is an important but unknown factor which may vary both regionally and through time. These complications highlight the importance of accurate reporting of all taken tests, as accurate estimates of LFT uptake are integral in understanding their impact.

While our model considers a time period before the Delta variant (B.1.617.2) dominated infections in the UK ^49^, and we do not explicitly consider the Delta variant in the model, we can nevertheless consider the implications of our work in the context of new, more transmissible variants. With *R*_*school*_ approximately equal to 1 in mid-May 2021, more transmissible variants such as the Delta variant (which has been estimated as 60% more transmissible ^59^) could tip the balance of transmission within secondary schools, increasing *R*_*school*_ substantially above 1. Further, our model considers a time-period where stringent within-school distancing measures were implemented. Prior studies have demonstrated that within-school measures can be effective in mitigating within-school transmission ^17,18^, and high attack rates in schools have been observed when such measures have not been in place ^60^. Any relaxation of within-school distancing measures would likely result in further increases in within-school transmission and hence *R*_*school*_. Further research into the impact of within-school control measures ^30,61^ should continue. Because of these factors, far higher uptake of LFT testing may be necessary to mitigate within-school infections in the future, especially to offset the impact of 60% extra transmission associated with the Delta variant. A range of socioeconomic factors impact LFT uptake, including the fear of loss of income that could result from a household required to self-isolate ^23^. Policy makers should therefore consider practical strategies that may incentivise or increase uptake of LFTs, especially for any future strategy that does not include the isolation of close contacts. For example, pupils could be expected to present a negative LFT test upon attending school, or pupils participating in serial contact testing could be tested within-school to encourage high levels of uptake. Dialogue between all relevant stakeholders, including pupils, parents, and staff, is crucial in the formulation of practical and effective policies that maximise participation.

Our analyses have considered the impact of LFTs in the context of secondary schools. In England, practical considerations have dictated that mass testing has not been extended to primary school children. If the isolation of close contacts is to be halted in primary schools, this raises the question of what control measures are implemented instead, whether they will be sufficient to control within-school transmission, and whether they are practical to implement for that age range.

Continued research into transmission within schools will be vital going forwards, both in understanding the role of children in transmission of SARS-CoV-2, and in designing and assessing appropriate school-level control policies. Previous studies undertaken prior to the COVID-19 pandemic have attempted to record contact mixing patterns within schools ^62–64^, however the implementation of rigid social distancing measures within schools mean that such studies are not of direct use in the context of COVID-19. The CoMix study has surveyed social contacts in the UK during the COVID-19 pandemic and has been used to infer age-dependent mixing matrices ^65^, though is not directly informative of contact structure within schools specifically. In this study we find that, once the model is calibrated to fit the available data, the relative levels of mixing assumed within schools has little bearing on the impact of control measures. An understanding of whether this holds in general, and a deeper understanding of the interplay between contact network structure within schools and the success of COVID-19 control measures, would be an important contribution going forward.

To conclude, through the use of a stochastic individual-based model, fitted to a wide range of relevant data, we demonstrate that twice weekly lateral flow testing has reduced within-school transmission of SARS-CoV-2 in England since its introduction to secondary school pupils, keeping *R*_*school*_ below 1 from March to May 2021. Yet, our results also indicate the delicate situation regarding transmission within secondary schools. With *R*_*school*_ only just below 1, increases in within-school transmission, either because of more transmissible variants or because of a relaxation of within-school distancing measures, are expected to result in within-school epidemics. We have shown the potential of serial contact testing alongside twice weekly mass testing to control within-school transmission while minimising the disruption caused by pupil absences, and the increased effectiveness of mass testing strategies at higher LFT uptake. Accordingly, our findings suggest that alternative strategies to the isolation of all close contacts are worth consideration, and that strategies involving LFTs may strike a balance between controlling within-school transmission and reducing pupil absences.

## Availability of data and materials

Data from the Department for Education Educational Settings database were supplied after anonymisation under strict data protection protocols agreed between the University of Warwick and the Department for Education in the UK. The ethics of the use of these data for these purposes was agreed by the Department for Education with the Government’s SPI-M(O)/SAGE committees.

Public Health England (PHE) collected data in a centralised database, which included details on the type of test and results. PHE provided anonymised data to contributors of the Scientific Pandemic Influenza Group on Modelling (SPI-M) as part of the COVID-19 response under a data sharing agreement between PHE and the authors’ institutions.

All other data sources used within this parameterise the study are publicly available and stated within the main manuscript.

Code for the model and model fitting is available at: https://github.com/tsleng93/SchoolReopeningStrategies

## Data Availability

Data from the Department for Education Educational Settings database were supplied after anonymisation under strict data protection protocols agreed between the University of Warwick and the Department for
Education in the UK. The ethics of the use of these data for these purposes was agreed by the Department for Education with the Government's SPI-M(O)/SAGE committees.
Public Health England (PHE) collected data in a centralised database, which included details on the type of test and results. PHE provided anonymised data to contributors of the Scientific Pandemic Influenza Group on Modelling (SPI-M) as part of the COVID-19 response under a data sharing agreement between
PHE and the authors' institutions.
All other data sources used within this parameterise the study are publicly available and stated within the main manuscript.
Code for the model and model fitting is available at: https://github.com/tsleng93/SchoolReopeningStrategies

## Acknowledgements

This work has been supported by the Engineering and Physical Sciences Research Council through the MathSys CDT [grant numbers EP/S022244/1 and EP/L015374/1];and by the Medical Research Council through the COVID-19 Rapid Response Rolling Call [grant number MR/V009761/1]. TL, MJK, LD and MJT were supported by Medical Research Council through the JUNIPER modelling consortium [grant number MR/V038613/1]. The funders had no role in study design, data collection and analysis, decision to publish, or preparation of the manuscript.

We thank Nick Gent at Public Health England for providing access to the data, and thank Ellen Brooks-Pollock for comments on an early draft.

## Supporting information items

### S1 Constructing modelled secondary schools

We derived from the Department for Education: Educational Setting Status data ^35^ the number of pupils attending a given modelled secondary school, the LTLA of a modelled secondary school, and close contact group sizes for September-December 2020 and for March-May 2021. The number of pupils for each secondary school within the data was taken as the median reported number of pupils on roll from September to December 2020; 2979 secondary schools report such data over this period. Whether a school included a sixth form was also specified – we assumed schools that had a sixth form to be comprised of seven year groups, while schools without a sixth form we assumed to be comprised of five year groups.

While the size of groups schools send home may vary through time and may be context dependent, we can use absences data from each term to obtain a proxy for the number of close contacts schools are sending home upon identification of a positive case. Specifically, we considered the size of groups different schools sent home after one case by finding the median number of COVID-19 related absences for each school, given there was one confirmed COVID-19 positive pupil (and no confirmed COVID-19 positive teachers). We found close contact group sizes for September-December 2020 and March-May 2021 separately, as schools in general isolated smaller groups of pupils in the latter term. Close contact group sizes for schools that did not report one confirmed COVID-19 positive pupil in each term were sampled from the distribution of reported close contact sizes (as a proportion of the school population). We adjusted school sizes, year group sizes, and close contact group sizes for both terms such that year group sizes were a divisor of school sizes, and close contact group sizes for both terms were a divisor of year group sizes. Doing so, we obtained a population of 2979 modelled schools with qualitatively similar population sizes and that implemented close contact isolation policies as reported in Department for Education data.

**Figure S1:**
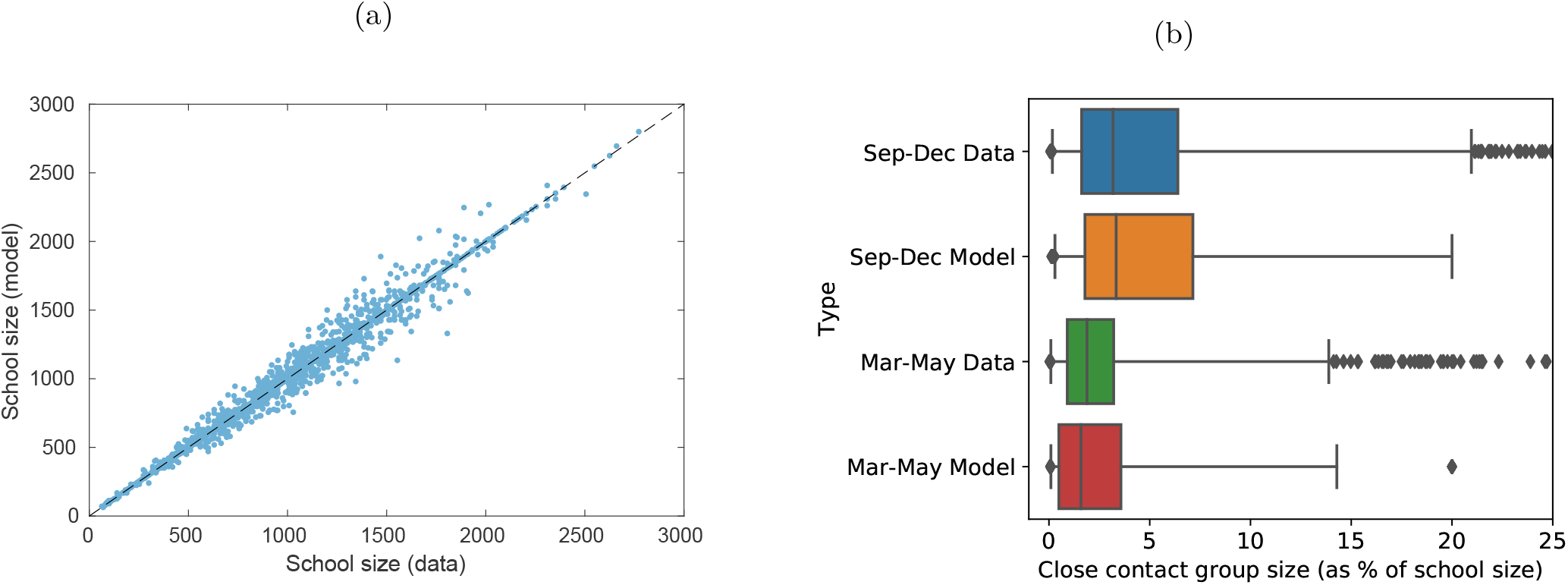
Comparing modelled secondary schools from absences data. (a) A scatter plot comparing the size of secondary schools obtained from Department for Education: Education Setting Status data, taken as the median reported number of pupils on roll from September to December 2020, against modelled secondary school sizes, after adjusting data such that close contact group sizes are divisors of year group sizes and year group sizes are divisors of school sizes. (b) Box plots comparing close contact group sizes inferred from absences data and close contact group sizes used within the model, as a percentage of school size across both terms. Whiskers refer to 2.5% and 97.5% percentiles. Close contact group sizes above 25% of the size of schools are omitted from the graph for box plots corresponding to data for both periods.

### S2 Transmission dynamics in detail

#### S2.1 Within-school transmission

As in our previous study ^27^, infected pupils attending school (i.e. those not isolating and it being a school day) transmitted infection to other pupils within their year group with a probability dependent on the time elapsed since their infection. Specifically, we assumed that the relative probability of transmission since the day of infection is given by a Gamma distribution (Γ_*I*_ (*d*)) with shape 5.62 and scale 0.98^41^, derived from data from known source-recipient pairs ^42^, with an assumed incubation period distribution (Gamma distributed with shape 5.807 and scale 0.948^43^) under the assumption that the generation time and incubation period are independent. After 14 days, infected individuals recover with immunity that persists over the course of the simulation. Infections are modelled from the 24th August 2020 (a week before schools reopened) until 21st May 2021. We assumed that pupils did not attend school, and therefore did not transmit infection within school, from the 24th-28th August 2020, from 26th-30th October 2020, and from 20th December 2020 - 5th March 2021, corresponding to periods when schools were either on holiday or not fully reopen due to lockdown measures. Additionally, pupils also did not attend school for two weeks out of the three from 29th March 2021 - 16th April 2021, with the specific weeks not attending school dependent on that school’s LTLA.

The initial level of transmission within a particular school *s, K*_*s*_ was drawn from a lognormal distribution: *K*_*s*_ *∼* LogNormal 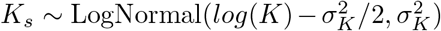, with the parameters *K* and *σ*_*K*_ determined via the fitting procedure, with *K* representing the mean value of *K*_*s*_ across schools. *K*_*s*_ defined the expected number of secondary cases from an infected symptomatic pupil in a particular school, assuming a fully susceptible school population, that pupils attend school each day of their infection, and that the impact of depletion of susceptibles is negligible. The model assumes that asymptomatic individuals are less infectious than symptomatic pupils by a factor *a*, 0 *≤a≤* 1 i.e. if a symptomatic pupil is expected to infect *K*_*s*_ other pupils over the course of their infection, an asymptomatic pupil is expected to infect *a ×K*_*s*_ other pupils. We define the function *A*_*i*_ to be such that *A*_*i*_ = 1 if a pupil will develop symptoms, otherwise *A*_*i*_ = *a*.

The level of within-school transmission on day *d* is also impacted by the proportion of cases within a school’s LTLA that are of the B.1.1.7 variant. This proportion is obtained by fitting a sigmoidal curve to the proportion of sequenced tests within an LTLA that return an S-gene negative result (Figure S2, a reliable genomic indicator of the B.1.1.7 variant. This proportion then scales the level of within-school transmission, i.e. if the new cases in an LTLA are 50% B.1.1.7 variant, then the increase in a pupil’s within-school transmission is 50% of that as if all new cases within that LTLA were of the B.1.1.7 variant. We let *V*_*LT LA*_(*d*) denote the scaling factor increase in within-school transmission resulting implied by the proportion of this variant in an LTLA on day *d*, with *V*_*LT LA*_(*d*) = *v*, 1 *≤v≤* 3 when all new cases are of the B.1.1.7 variant.

**Figure S2:**
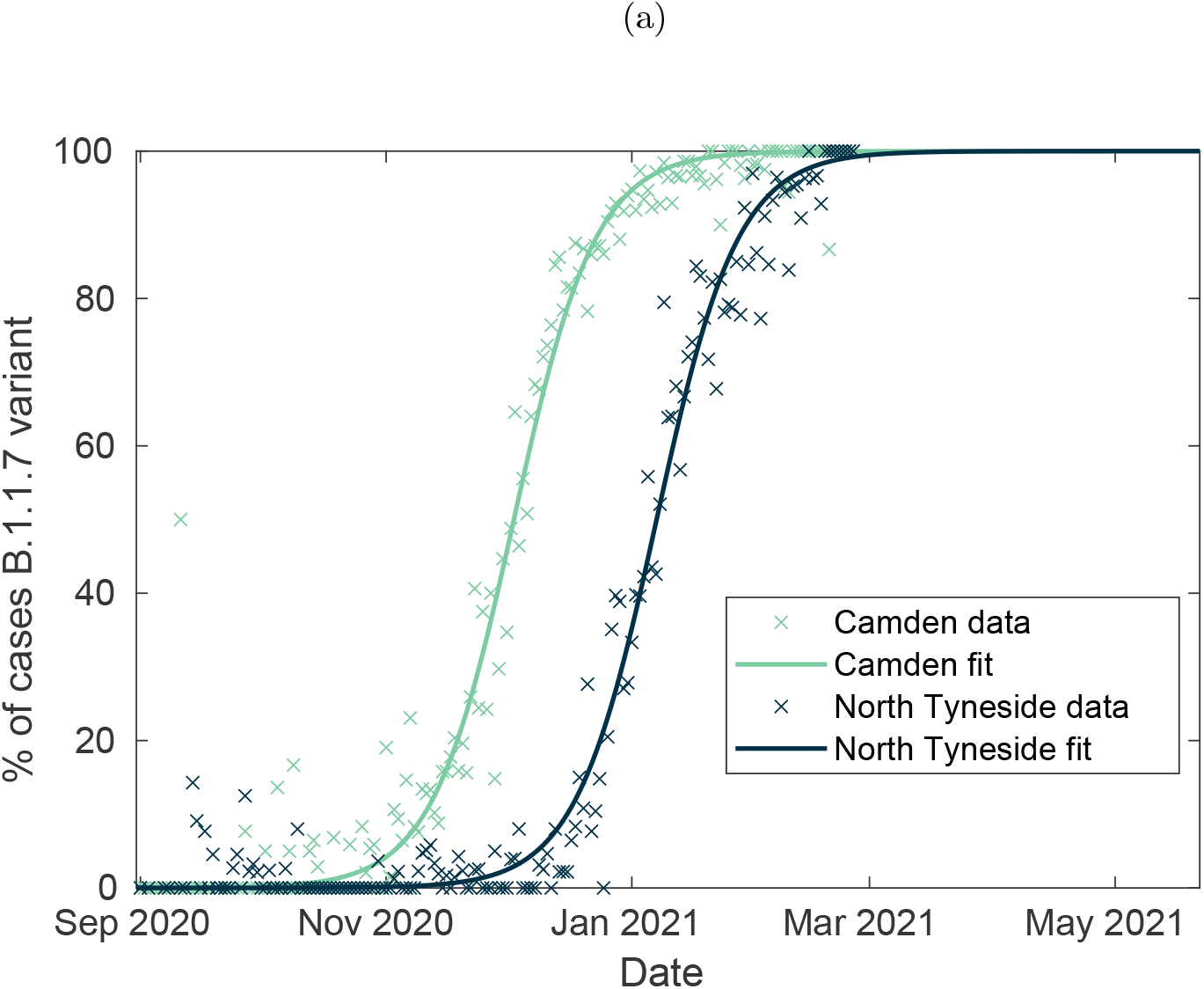
Approximating the relative frequency of B.1.1.7 variant cases. The proportion of sequenced tests (cross markers) for each LTLA that returns an S-gene negative increases approximately sigmoidally through time. As an S-gene negative result is a reliable genomic marker of the B.1.1.7 variant, we approximate the percentage of cases of the B.1.1.7 variant through time by fitting a sigmoidal curve to this data (solid lines). This data, and fitted sigmoidal curves, are plotted for two exemplar LTLAs.

We also introduce *F* (*d*), a function scaling up within-school transmission for all schools by a constant factor *f*, 1*≤ f ≤*2 from 2nd November 2020 onwards, i.e. after the first half-term of schools reopening. This parameter is to account for the impact of reduced adherence to within-school control measures as the school term progresses, after initially very high adherence to such measures, and is also determined via the fitting procedure.

For a pupil *i* infected on day *d*_0_, attending a school *s* situated in a particular LTLA on day *d*, the infectiousness of individual *i* on day *d* is given by:

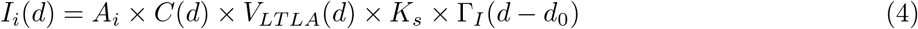

#### S2.2 External infection

All pupils from a school *s* in a given LTLA who were not isolating had a probability of external infection each day *d*, proportional to the proportion of the LTLA’s population who tested PCR positive on day *d* (which we denote 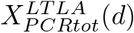), obtained from Pillar 2 data. For each school *s*, 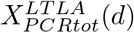 is scaled by a factor 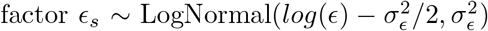, with parameters *E* and *σ* determined via the fitting procedure, and with *E* representing the mean scaling factor of *E*_*s*_ across schools.

The probability of external infection for pupils in schools in rural LTLAs is scaled down by a factor *r*, 0 *< r ≤*1. We introduce the function *R*_*LT LA*_, which equals 1 for urban LTLAs and *r* for rural LTLAs. The probability of external infection for pupils is scaled up during school holidays and school closures by a factor *h*, 1*≤ h ≤*2, because during such periods pupils will spend more time with non-school contacts, hence increasing their risk of community infection. We introduce the function *H*(*d*) which equals 1 during term time and *h* during school holidays or closures. For a susceptible pupil *j*, who attends school *s* situated in a particular LTLA, the probability that they are infected via community infection is given by *E*_*s*_(*d*):

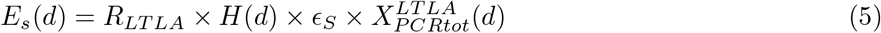

Pupil *j*’s total probability of infection on day *d* is given by *β*_*j*_(*d*) + *E*_*s*_(*d*), with *β*_*j*_(*d*) as defined in Equation (1)

### S3 LFT uptake

This section describes how mass testing via LFTs, and realistic levels of LFT uptake per LTLA, are implemented. Within the model, twice weekly mass testing of secondary school pupils is implemented from the 1st March 2021 until the end of the simulation, in line with the introduction of mass testing prior to secondary schools reopening on the 8th March 2021. Each pupil was assigned two days of the week they are scheduled to take their tests - taking their first test on Sunday, Monday, Tuesday, or Wednesday, and taking their second test three days after. The probability of a pupil scheduled to take a test *p* was adjusted to match daily LFT uptake in secondary school aged pupils within that school’s LTLA.

Daily LFT uptake per LTLA was taken as the proportion of 10-19 year olds in that LTLA who recorded either a positive or negative LFT on that day. We used the broader age range of 10-19 year olds here due to the negative Pillar 2 data only being available in five year age bands. This is likely an underestimate, for two reasons. Firstly, a considerable proportion of negative home tests remain unrecorded. To account, we determine the level of underreporting, *u*, 1*≤ u ≤* 4, via the fitting procedure. The number of negative LFTs per LTLA is scaled up by the factor of *u* from 14th March 2021, i.e. after tests shifted from being at school to being taken at home. We assume that *u* remains constant over time and is constant across LTLAs, though in reality *u* may wain over time and vary by LTLA. Secondly, the age range considered includes some ages not attending secondary school (who are not expected to take LFT tests), and consequently testing uptake is likely concentrated in a smaller age range. While this is not accounted for explicitly, the scaling up via underreporting counteracts this underestimation. LFT uptake obtained from data and modelled uptake after accounting for underreporting is shown in Supporting Figure S3a.

Estimates of LFT uptake per LTLA over the term are taken as the mean daily LFT uptake from 8th March 2021 until 23rd May 2021, divided by 2/7 - as 2/7ths of pupils taking a test every day would correspond to full uptake of twice weekly mass testing (assuming that the day of first tests and day of second tests are distinct and random). There is considerable heterogeneity in LFT uptake between LTLAs, shown in Supporting Figure S3b.

**Figure S3:**
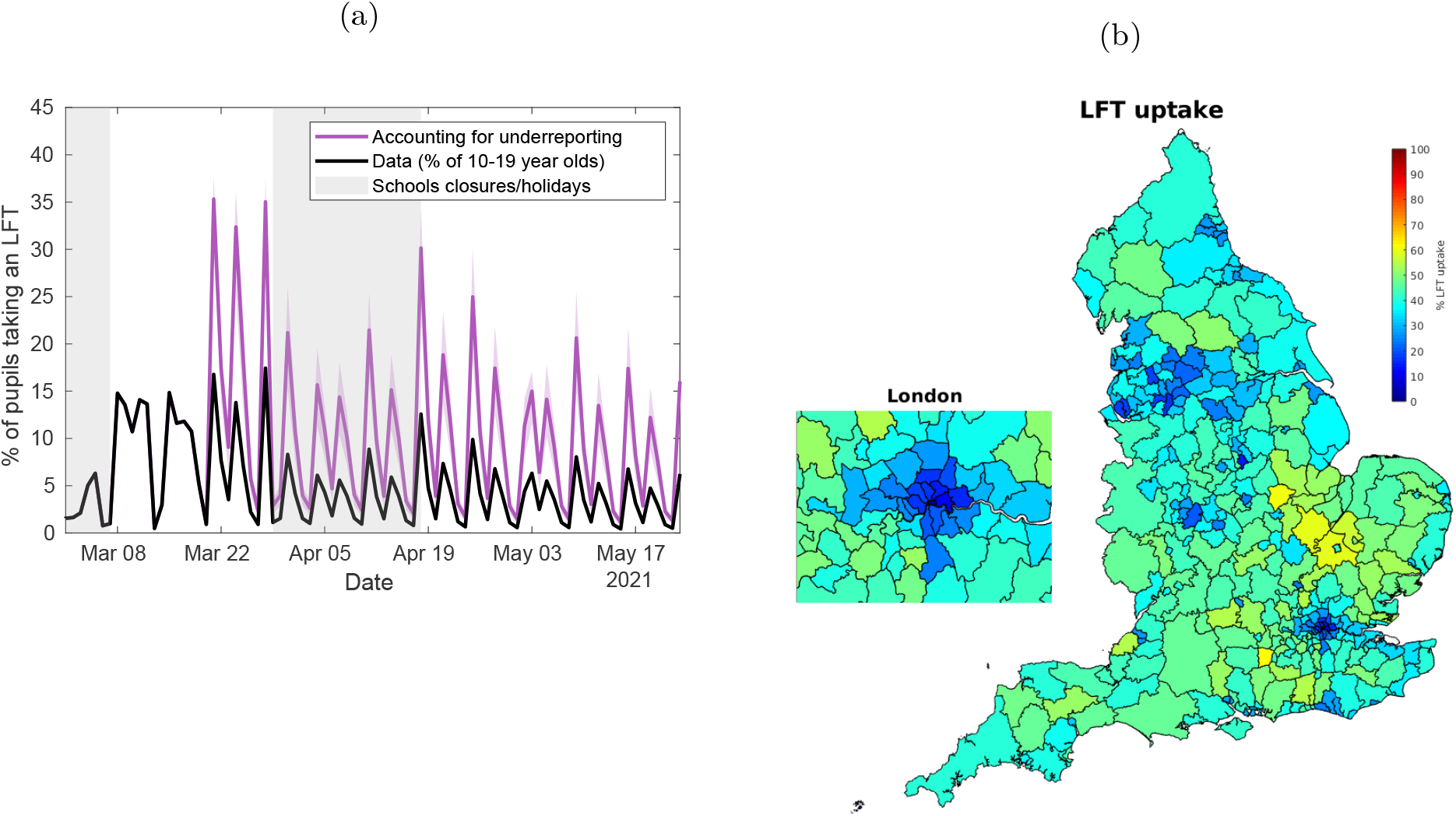
Modelling LFT uptake. (a) We plot time-series of the percentage of 10-19 year olds in England who report either a positive or negative LFT result each day (black line), and of the modelled percentage of pupils who taken a LFT each day after accounting for underreporting of home tests from 21st March 2020 (purple line). Model results are from 100 simulations of 2979 secondary schools, each with a distinct parameter set sample from the posterior distribution, with solid purple line corresponding to the mean estimate, and the shaded purple interval representing the 95% prediction interval. (b) A map segregated into LTLAs, where each LTLA is shaded according to modelled percentage uptake of LFTs from 8th March-16th May 2021, taken as the mean daily LFT uptake within that LTLA from 8th March 2021 until 16th May 2021, divided by 2/7.

### S4 Model fits

In this section, Table S1 describes and explains each model parameter, the prior distribution assumed for each parameter, and the 95% credible interval of the posterior distribution for each parameter. Figure S4 displays density plots of a sample of 100 parameter sets obtained from the posterior distribution. Figure S5 demonstrates the improvement in fit obtained in successive generations via the fitting process, plotting the median log-likelihood obtained per generation.

**Table S1:**
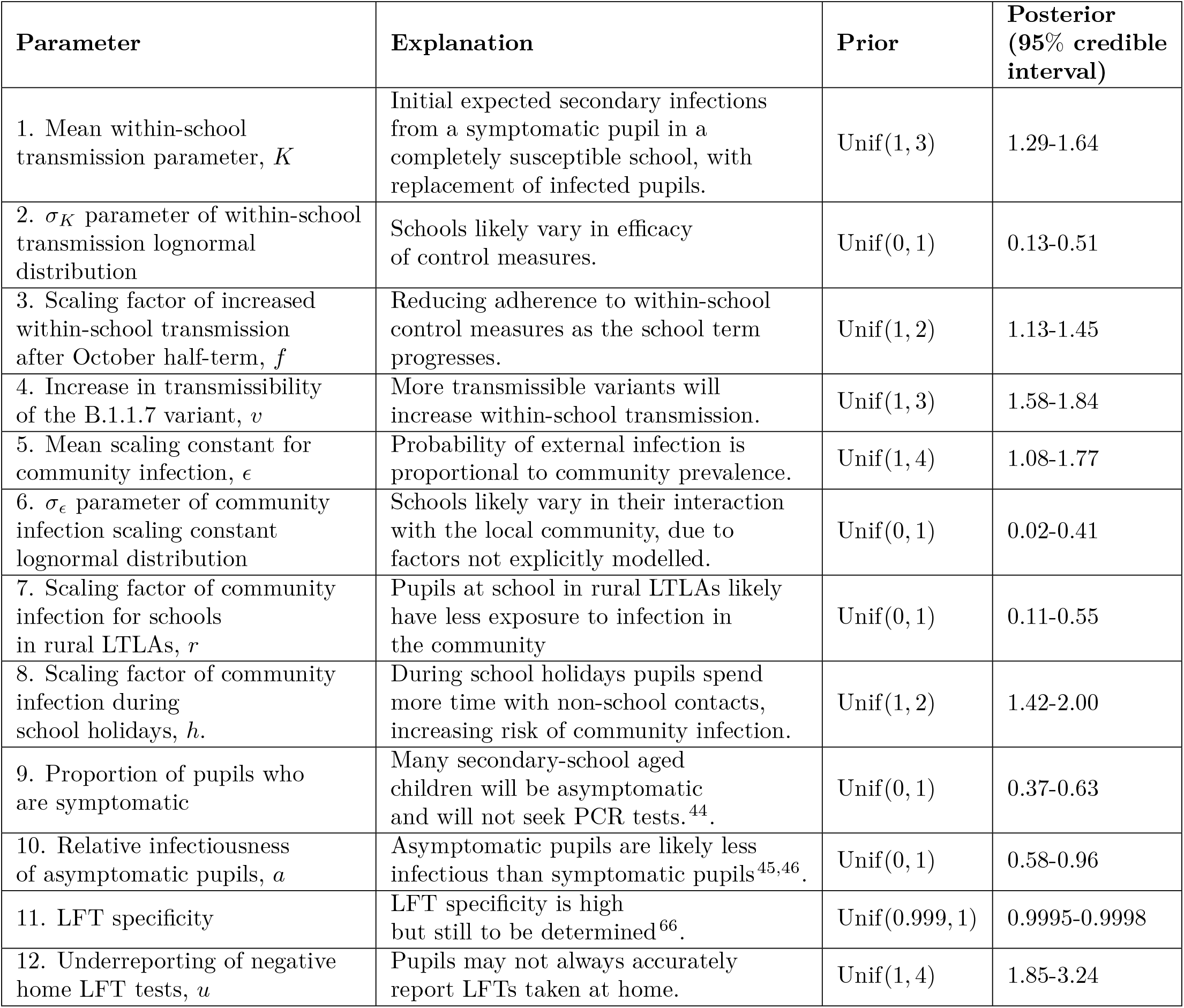
Explanation of model parameters, assumed prior distributions, and posterior ranges. All posterior ranges are specified to two decimal places, with the exception of LFT specificity that we specify to four decimal places to reflect the relative narrowness of the prior.

**Figure S4:**
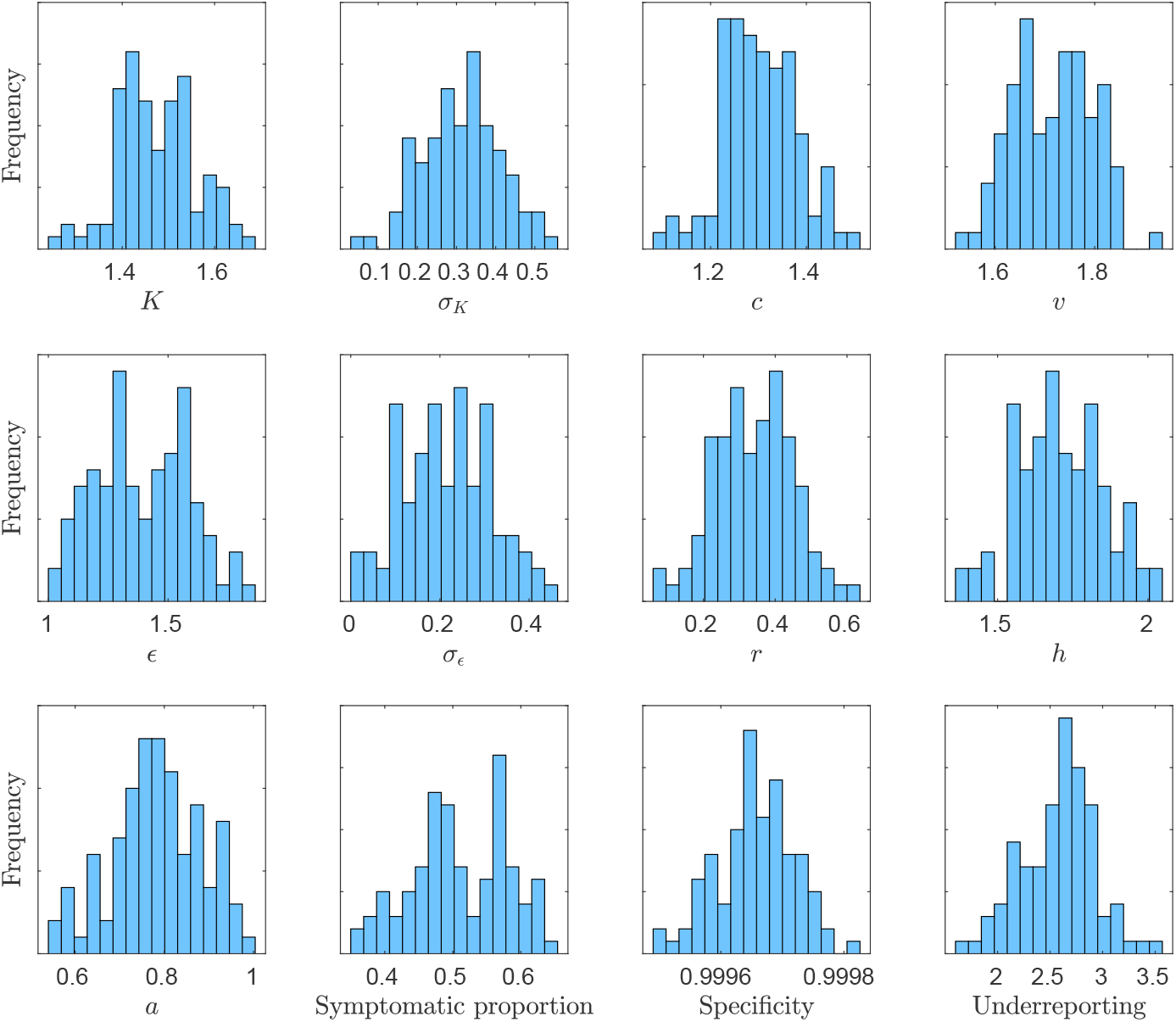
Posterior distribution density plots. Plotted are posterior parameter density plots for each of the parameters fitted in the model via an ABC approach. Density plots of 100 parameter sets sampled from the posterior distribution. We observe the density of each parameter to be approximately normally distributed.

**Figure S5:**
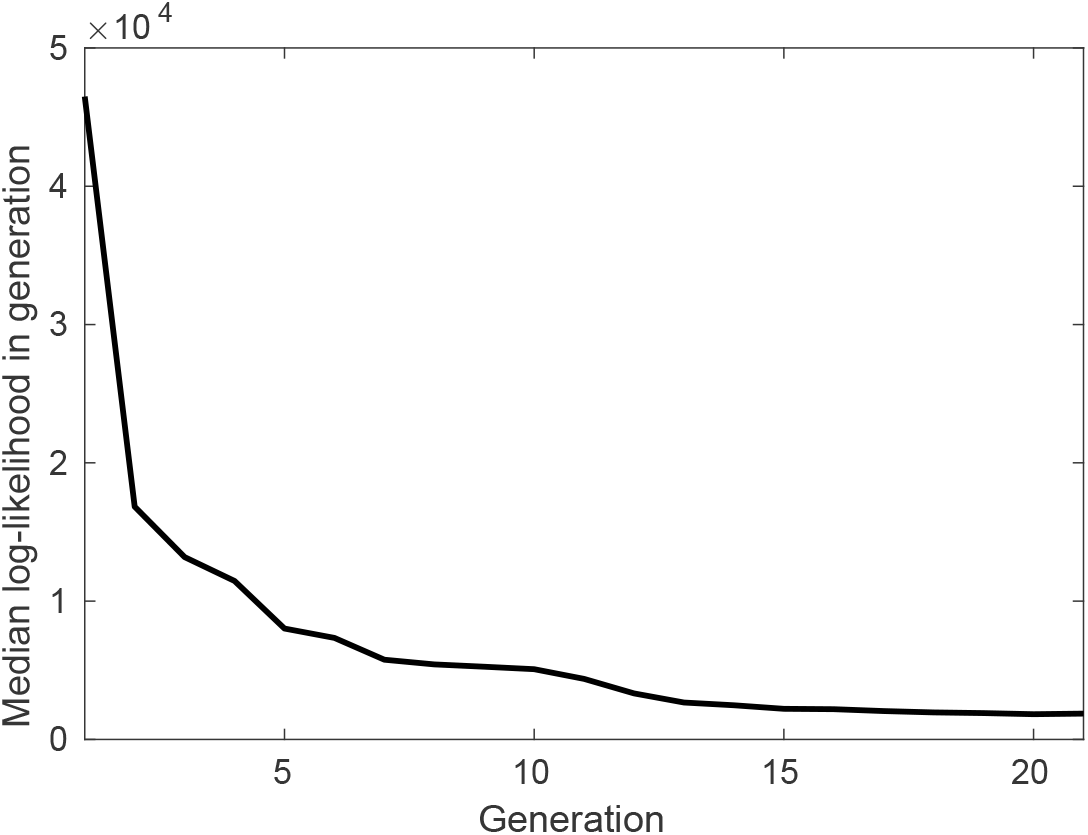
Model fitting by generation. We display the median log likelihood per generation of 100 parameter sets explored in each generation. For each parameter set, we simulated epidemics in 100 randomly sampled schools for the initial ten generations, to navigate the parameter space quickly. After the tenth generation, we simulated epidemics in 1000 randomly sampled schools. We proceeded to a further generation until the median log likelihood per generation no longer decreased.

### S5 Comparing the model against absences data

**Figure S6:**
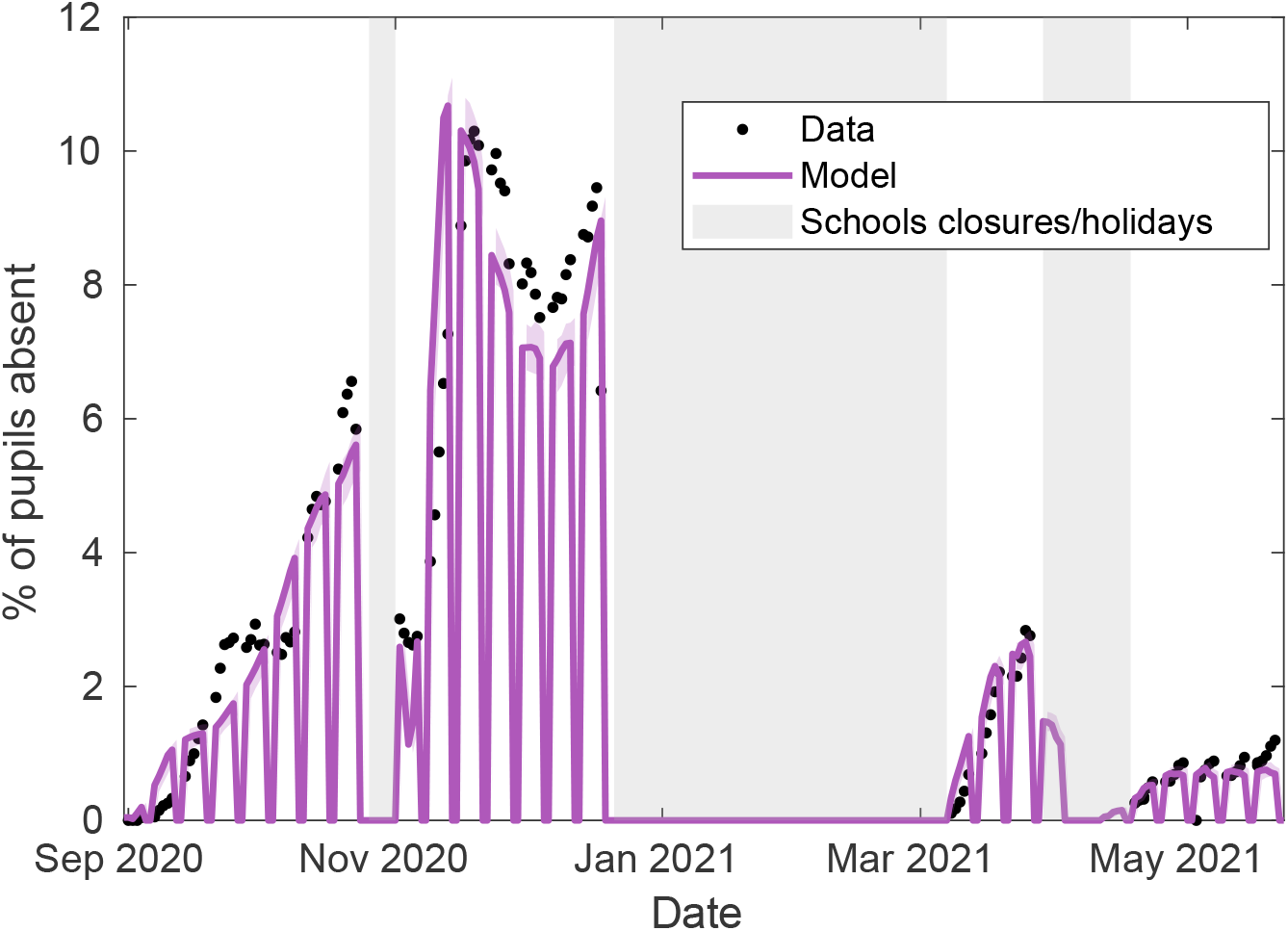
Comparing modelled absences against within-school COVID-19 related absences data. The time-series of the modelled percentage of pupils absences (purple line) is compared to the observed percentage of pupils absent due to a confirmed case or a suspected case of COVID-19, or due to being told to isolate due to potential contact with a case of COVID-19 from inside their educational setting, for the 2979 secondary schools that recorded this data. Shaded intervals represent 95% prediction intervals.

### S6 Comparing the model against data by region and by LTLA

**Figure S7:**
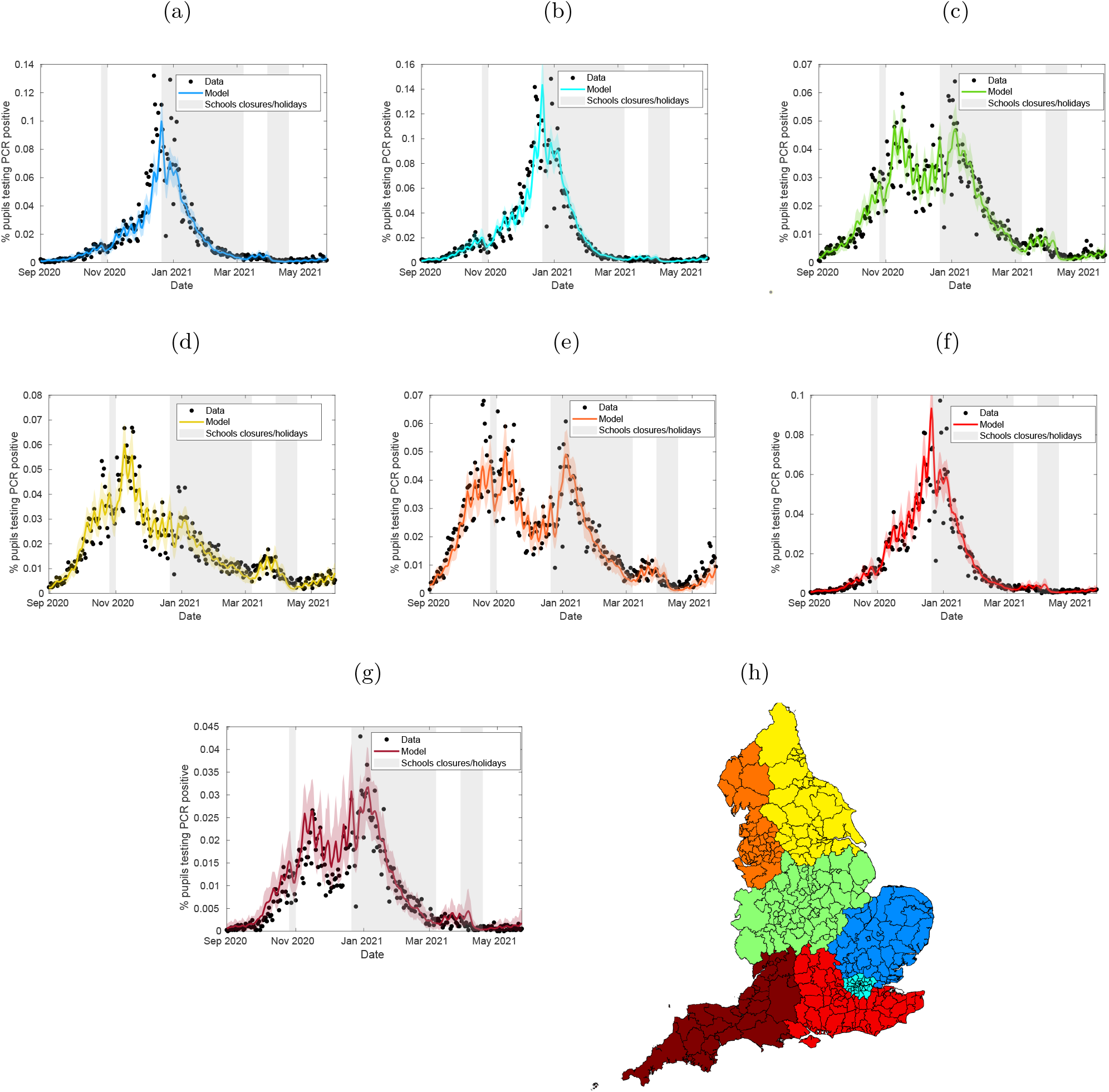
Comparing the fitted model against PCR testing data by region. The above plots compare the percentage of 11-16 year olds who test PCR positive regionally (excluding confirmatory PCR tests) each day from 1st September 2020 to 21st May 2021 against the modelled percentage of pupils testing PCR positive through time, for (a) East of England, (b) Greater London, (c), the Midlands, (d) the North-West, (e), the North-East, (f) the South-East, and (g) the South-West, with (h) showing the LTLAs that aggregated form each region, with colours corresponding to the time-series plots. Plots above were obtained from 100 simulations of 2979 secondary schools, each with a distinct parameter set sample from the posterior distribution. In all panels, solid lines correspond to mean temporal profiles, while shaded ribbons represent 95% prediction intervals.

**Figure S8:**
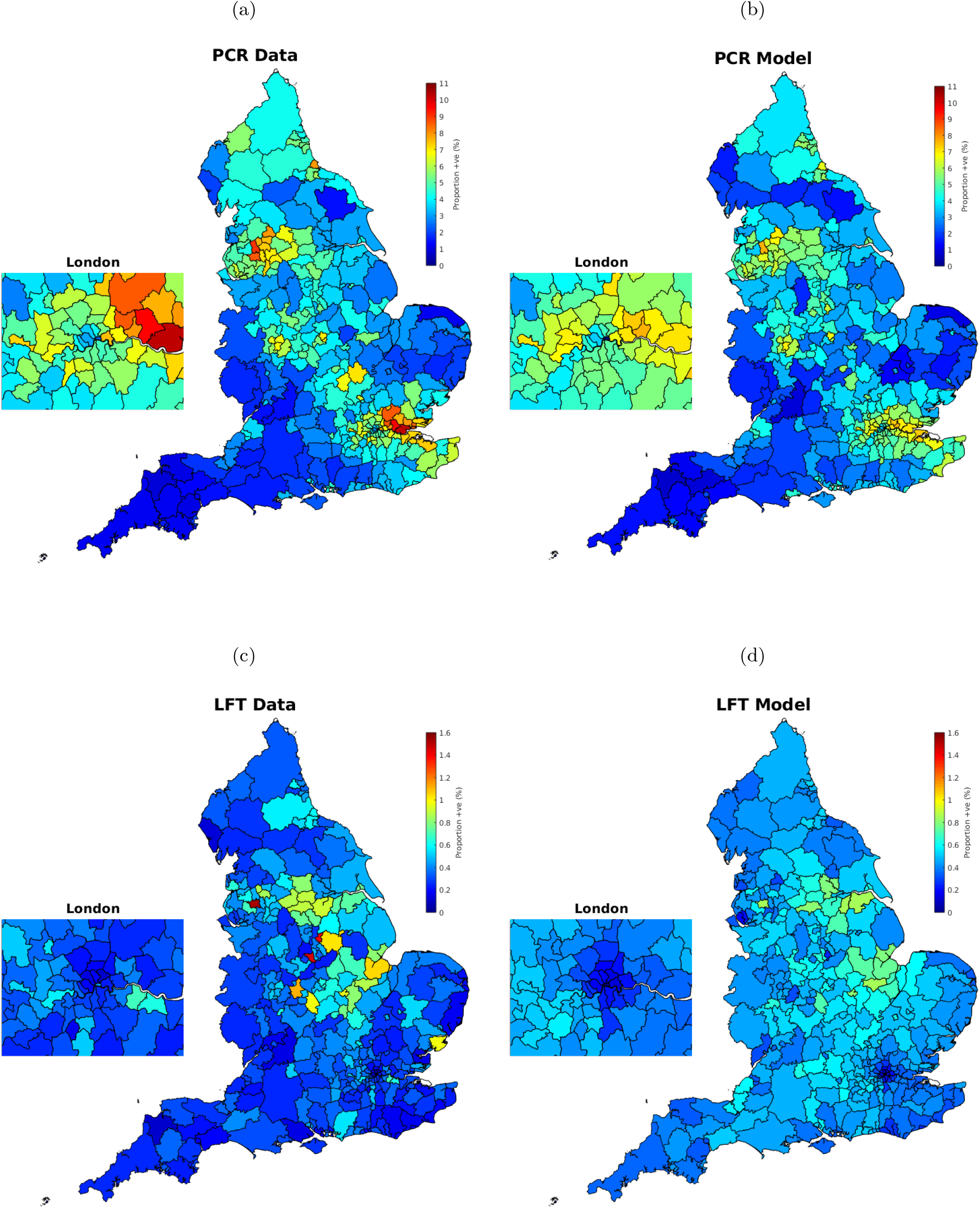
Comparing the fitted model against testing data by LTLA. Each LTLA is shaded to signify (a) the proportion of 11-16 year olds and (b) the proportion of modelled pupils from the fitted model who have tested positive to a PCR test (excluding confirmatory PCR tests) from 1st September 2020 until 21st May 2021, shaded according to the same colour range. Similarly, Each LTLA is shaded to signify (c) the proportion of 11-16 year olds and (d) the proportion of modelled pupils from the fitted model who have tested positive to an LFT test from 8th March 2021 until 21st May 2021, shaded according to the same colour range. Results were obtained from 100 simulations of 2979 secondary schools, each with a distinct parameter set sample from the posterior distribution.

### S7 Assuming greater relative levels of within-year and between-year mixing

This section contains analogous figures to Figures 2 to 4, under alternative within-school mixing assumptions. In the main analyses presented in the main body of the paper, we assumed that pupils interacted with their close contacts at a much higher rate than other members of their year group, and where year group bubbles have been in general very effective, setting *α*_1_ = 0.1 and *α*_2_ = 0.01. To assess the sensitivity of our results to these mixing assumptions, we also fit our model under the assumption that pupils within their year-group mix randomly, setting *α*_1_ = 1 and that there is a much higher-level of interaction between year groups, setting *α*_2_ = 0.1. It is important to note that the differing mixing assumptions between this section and the main paper only effects *who* an infected pupil will infect, rather than *how many*. These results should not be interpreted as justifying a relaxation of within-school distancing measures implemented, as doing so would not only change the mixing patterns within a school, but would also likely increase the number of infectious contacts pupils make.

Assuming *α*_1_ = 1, *α*_2_ = 0.1, we fitted the model via the ABC procedure outlined in Section 2.3. The fitted model matches well to both temporal PCR and LFT positivity data (Figures S9a and S9b). This alternative fitted model provided a better fit to the distribution of peak confirmed cases from September-December 2020 than the fitted model in the main analysis Figure S9c, but overestimated the proportion of schools with a low peak number of confirmed cases from March-May 2021 to a greater extent than the main analysis fitted model Figure S9d.

Qualitatively similar trends emerged in within-school transmission and *R*_*school*_, despite the differing assumptions regarding within school mixing Figure S10. The proportion of cases occurring within school increased through time, accounting for 37% (95% credible interval: 28-47%) of all new infections in the September-October half term, 69%: (95% credible interval: 61-75%) of all new infections in the November-December half term, and 74% (95% credible interval: 64-78%) of infections since schools reopened from 8th March 2021 until 21st May 2021.

As in the main analyses, the introduction of mass testing alongside isolation of close contacts substantially reduced incidence from 8th March-22nd May 2021 compared to a policy of isolation of close contacts only, and kept *R*_*school*_ below 1 over the same period. Under these alternative mixing assumptions *R*_*school*_ values obtained for each scenario were qualitatively similar to, though slightly lower to, *R*_*school*_ values obtained for their counterparts in the main analysis. As in the main analysis, a strategy of mass testing alone, or a strategy of serial contact testing alongside mass testing led to substantially lower levels of absences than either strategy involving the isolation of close contacts (Figure S11).

**Figure S9:**
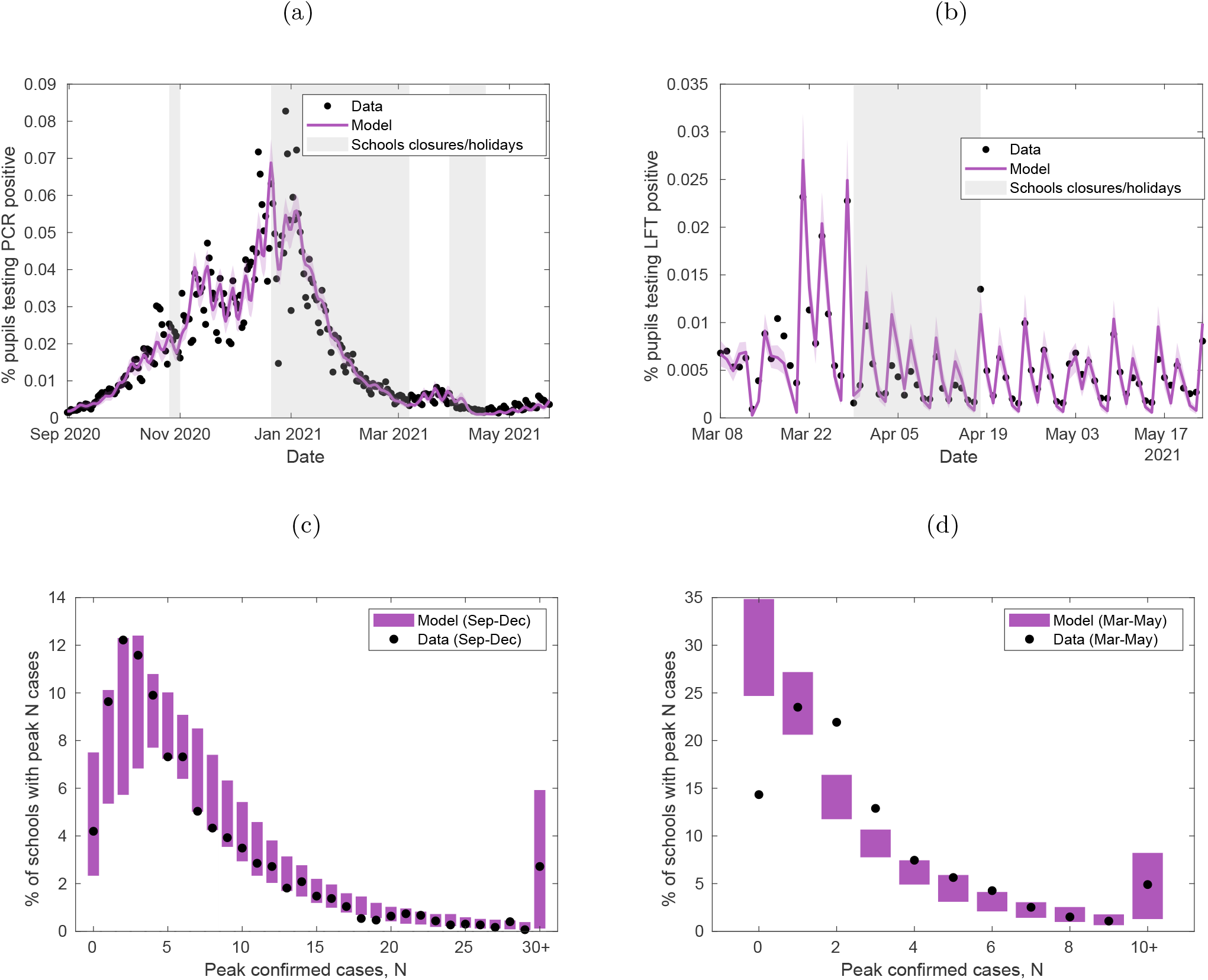
Fitting the model to testing and secondary school absences data, under different within-school mixing assumptions. Under the alternative within-school mixing assumptions *α*_1_ = 1, *α*_2_ = 0.1, the stochastic individual-based model is fitted to (a) the percentage of 11-16 year olds who test PCR positive (excluding confirmatory PCR tests) each day from 1st September 2020 to 21st May 2021, the percentage of 11-16 year olds who test LFT positive each day from 8th March 2021 to 21st May 2021. Shaded intervals around mean model traces (solid lines) represent 95% prediction intervals in all plots, while shaded grey regions represent time periods when schools were not fully open (either due to closures or school holidays). The model is also fitted to (c) the distribution of peak number of confirmed COVID-19 absences in secondary schools from 1st September 2020 to 18th December 2020, and (d) the distribution of peak number of confirmed COVID-19 absences in secondary schools from 8th March 2021 to 21st May 2021. Filled circles denote the data and shaded blocks the 95% prediction interval estimated from the model. The model provides a reasonable fit to both distributions, though underestimates the proportion of schools that have a low peak number of confirmed cases in (c), while overestimates the proportion of schools with a low peak number of confirmed cases in (d). The plots above show the mean values obtained from 100 simulations of 2979 secondary schools, each with a distinct parameter set sample from the posterior distribution.

**Figure S10:**
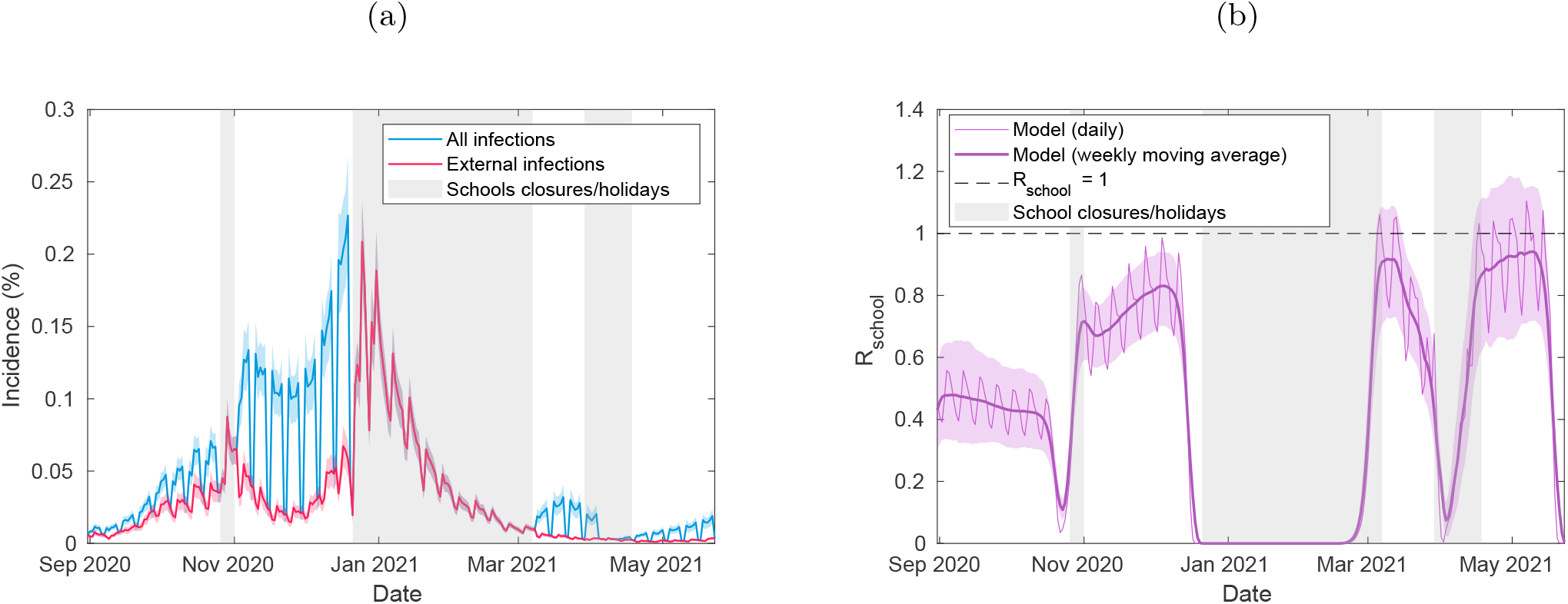
Incidence and *R*_*school*_ from the fitted model, under different within-school mixing assumptions. For the model fitted under the alternative within-school mixing assumptions *α*_1_ = 1, *α*_2_ = 0.1, we display time-series of (a) incidence among pupils, delineated into whether infections occur externally or within-school, and (b) *R*_*school*_ through time (thin line) alongside its seven-day moving average (thick line). The plots above were obtained from 100 simulations of 2979 secondary schools, each with a distinct parameter set sample from the posterior distribution. In all panels, solid lines correspond to mean temporal profiles, shaded ribbons represent 95% prediction intervals in all plots, while shaded grey regions represent time periods when schools were not fully reopen (either due to closures or school holidays).

**Figure S11:**
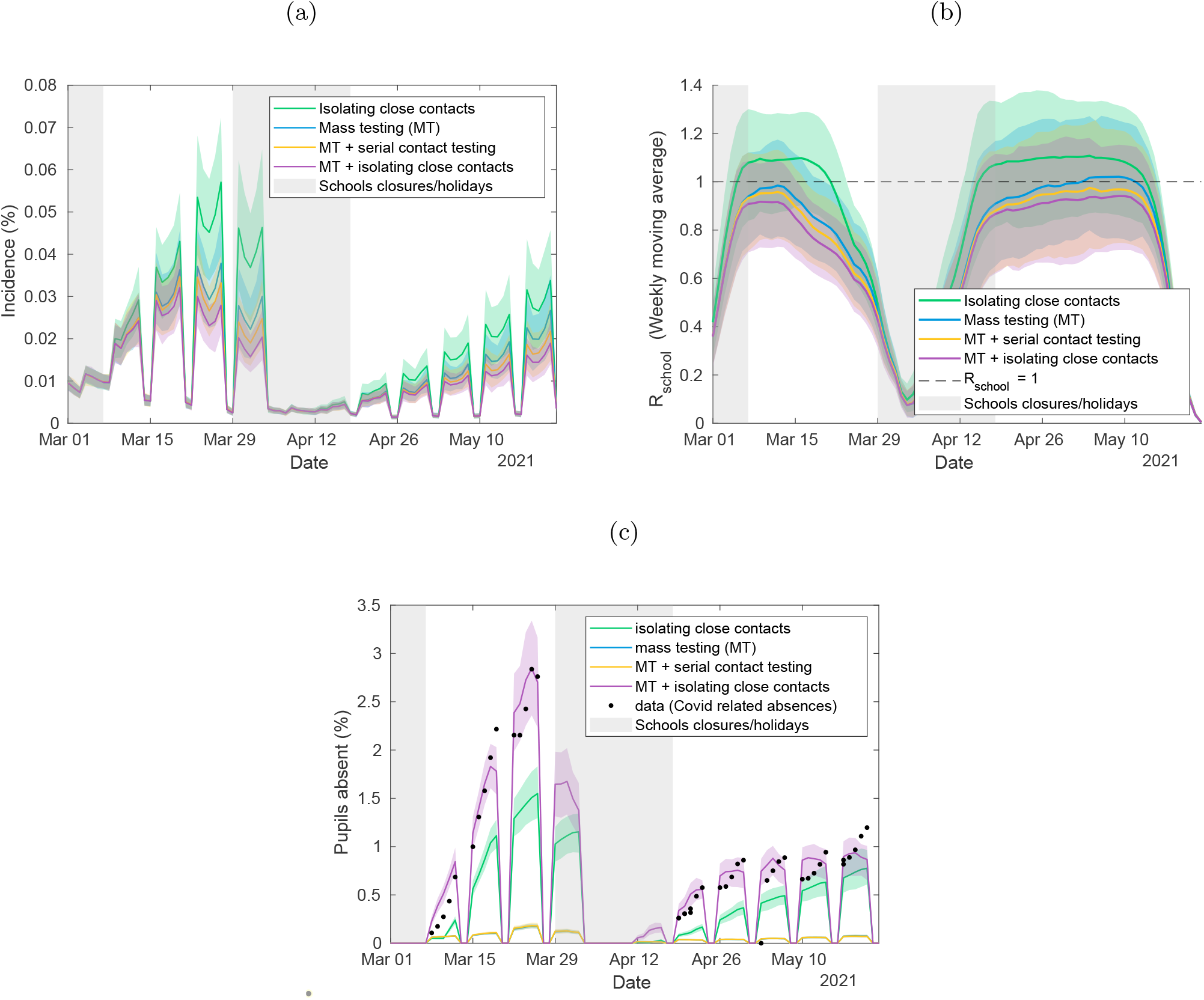
Quantifying the impact of LFTs on transmission and absences, and the potential impact of alternative strategies, under different within-school mixing assumptions. Time-series under different intervention strategies for the model fitted under the alternative within-school mixing assumptions *α*_1_ = 1, *α*_2_ = 0.1 of (a) incidence among pupils, (b) *R*_*school*_ within secondary schools, and (c) the percentage of pupils absent. We compare a policy of twice weekly mass testing and isolating close contacts (purple) to a strategy of isolating close contacts only (green), twice weekly mass testing only (blue), and twice weekly mass testing alongside serial contact testing (yellow). The plots above show the mean values obtained from 100 simulations of 2979 secondary schools, each with a distinct parameter set sample from the posterior distribution. In all panels, solid lines correspond to the mean estimate, shaded intervals represent 95% prediction intervals, while shaded grey regions represent time periods when schools were not fully reopen (either due to closures or school holidays). The data in figure 4c consists of the number of absences due to a confirmed case or a suspected case of COVID-19, and absences arising as a result of students told to isolate due to potential contact with a case of COVID-19 from inside their educational setting. This is taken from the 2979 secondary schools that recorded this data.

## References

[1] Gottfried MA. The detrimental effects of missing school: Evidence from urban siblings. American Journal of Education. 2011;117(2):147–182.

[2] Burgess S, Sievertsen HH. Schools, skills, and learning: The impact of COVID-19 on education. VoxEu org. 2020;1(2).

[3] DELVE. Balancing the Risks of Pupils Returning to Schools; 2021. Available from: https://rs-delve.github.io/reports/2020/07/24/balancing-the-risk-of-pupils-returning-to-schools.html.

[4] Centre for Education Policy and Equalising Opportunities. Briefing Note: School Absences and Pupil Achievement; 2021. Available from: https://repec-cepeo.ucl.ac.uk/cepeob/cepeobn1.pdf.

[5] Education Endowment Fund. School Closures Rapid Evidence Assessment; 2020. Available from: https://educationendowmentfoundation.org.uk/evidence-summaries/evidence-reviews/school-closures-rapid-evidence-assessment/.

[6] NHS Digital. Mental Health of Children and Young People in England, 2020; 2021. Available from: https://files.digital.nhs.uk/AF/AECD6B/mhcyp_2020_rep_v2.pdf.

[7] Garstang J, Debelle G, Anand I, Armstrong J, Botcher E, Chaplin H, et al. Effect of COVID-19 lockdown on child protection medical assessments: a retrospective observational study in Birmingham, UK. BMJ Open. 2020 sep;10(9):e042867.

[8] Office for National Statistics. Coronavirus (COVID-19) Infection Survey, UK: 7 July 2021; 2021. Available from: https://www.ons.gov.uk/peoplepopulationandcommunity/healthandsocialcare/conditionsanddiseases/bulletins/coronaviruscovid19infectionsurveyantibodyandvaccinationdatafortheuk/latest.

[9] Ludvigsson JF. Systematic review of COVID-19 in children shows milder cases and a better prognosis than adults. Acta Paediatrica, International Journal of Paediatrics. 2020;109(March):1088–1095.

[10] Barton M, Mehta K, Kumar K, Lu J, le Saux N, Sampson M, et al. COVID-19 Infection in Children: Estimating Pediatric Morbidity and Mortality. medRxiv preprint. 2020:1–26.

[11] NHS England. COVID-19 total announced deaths 28 January 2021.; 2021. Available from: https://www.england.nhs.uk/statistics/wp-content/uploads/sites/2/2021/01/COVID-19-total-announced-deaths-28-January-2021.xlsx.

[12] Buonsenso D, Munblit D, De Rose C, Sinatti D, Ricchiuto A, Carfi A, et al. Preliminary evidence on long COVID in children. MedRxiv. 2021.

[13] Buonsenso D, Espuny Pujol F, Munblit D, Mcfarland S, Simpson F. Clinical Characteristics, Activity Levels and Mental Health Problems in Children with Long COVID: A Survey of 510 Children. Preprints. 2021:2021030271.

[14] Mensah AA, Sinnathamby M, Zaidi A, Coughlan L, Simmons R, Ismail SA, et al. SARS-CoV-2 infections in children following the full re-opening of schools and the impact of national lockdown: Prospective, national observational cohort surveillance, July-December 2020, England. J Infect. 2021 apr;82(4):67–74. Available from: https://linkinghub.elsevier.com/retrieve/pii/S0163445321000931.

[15] Southall ER, Holmes A, Hill EM, Atkins BD, Leng T, Thompson RN, et al. An analysis of school absences in England during the Covid-19 pandemic. medRxiv. 2021:2021.02.10.21251484. Available from: http://medrxiv.org/content/early/2021/02/16/2021.02.10.21251484.abstract.

[16] Rozhnova G, van Dorp CH, Bruijning-Verhagen P, Bootsma MC, van de Wijgert JH, Bonten MJ, et al. Model-based evaluation of school-and non-school-related measures to control the COVID-19 pandemic. Nature Communications. 2021;12(1614).

[17] Zimmerman KO, Akinboyo IC, Brookhart MA, Boutzoukas AE, McGann KA, Smith MJ, et al. Incidence and secondary transmission of SARS-CoV-2 infections in schools. Pediatrics. 2021;147(4).

[18] Falk A, Benda A, Falk P, Steffen S, Wallace Z, Høeg TB. COVID-19 cases and transmission in 17 K–12 schools—Wood County, Wisconsin, August 31–November 29, 2020. Morbidity and Mortality Weekly Report. 2021;70(4):136.

[19] Lessler J, Grabowski MK, Grantz KH, Badillo-Goicoechea E, Metcalf CJE, Lupton-Smith C, et al. Household COVID-19 risk and in-person schooling. Science. 2021;372(6546):1092–1097.

[20] UK Government. Guidance for schools: coronavirus (COVID-19); 2020. Available from: https://www.gov.uk/government/collections/guidance-for-schools-coronavirus-covid-19.

[21] UK Government. Guidance: Face coverings in education; 2021. Available from: https://www.gov.uk/government/publications/face-coverings-in-education/face-coverings-in-education.

[22] Pickering S, Batra R, Merrick B, Snell LB, Nebbia G, Douthwaite S, et al. Comparative performance of SARS-CoV-2 lateral flow antigen tests and association with detection of infectious virus in clinical specimens: a single-centre laboratory evaluation study. The Lancet Microbe. 2021.

[23] University of Liverpool. Liverpool Covid-SMART Community Testing Pilot: Evaluation Report: 17 June 2021; 2021. Available from: https://www.liverpool.ac.uk/media/livacuk/research/research-themes/Liverpool_Community_Testing_Report,(5).pdf.

[24] Larremore DB, Wilder B, Lester E, Shehata S, Burke JM, Hay JA, et al. Test sensitivity is secondary to frequency and turnaround time for COVID-19 screening. Sci Adv. 2020 nov:eabd5393. Available from: http://advances.sciencemag.org/content/early/2020/11/20/sciadv.abd5393.1.abstract.

[25] Department for Health and Social Care (UK). ISRCTN18100261 Daily contact testing schools and colleges trial. ISRCTNregistry. 2021. Available from: https://doi.org/10.1186/ISRCTN18100261.

[26] UK Government. Position statement regarding daily contact testing in schools from PHE and NHS Test and Trace; 2021. Available from: https://www.gov.uk/government/publications/daily-contact-testing-in-schools-statement-from-phe-and-nhs-tt-about-next-steps/position-statement-regarding-daily-contact-testing-in-schools-from-phe-and-nhs-test-and-trace.

[27] Leng T, Hill EM, Thompson RN, Tildesley MJ, Keeling MJ, Dyson L. Assessing the impact of secondary school reopening strategies on within-school COVID-19 transmission and absences: a modelling study. medRxiv. 2021:2021.02.11.21251587. Available from: http://medrxiv.org/content/early/2021/02/12/2021.02.11.21251587.abstract.

[28] Kunzmann K, Lingjaerde C, Bird S, Richardson S. The ‘how’ matters: A simulation-based assessment of the potential contributions of LFD tests for school reopening in England. arXiv preprint 210302035. 2021.

[29] Asgary A, Cojocaru MG, Najafabadi MM, Wu J. Simulating preventative testing of SARS-CoV-2 in schools: policy implications. BMC Public Health. 2021;21(1):1–18.

[30] Panovska-Griffiths J, Stuart RM, Kerr C, Rosenfeld K, Mistry D, Waites W, et al. Modelling the impact of reopening schools in early 2021 in the presence of the new SARS-CoV-2 variant and with roll-out of vaccination against COVID-19. medRxiv. 2021:2021.02.07.21251287. Available from: http://medrxiv.org/content/early/2021/02/09/2021.02.07.21251287.abstract.

[31] Kaiser AK, Kretschmer D, Leszczensky L. Social network-based strategies for classroom size reduction can help limit outbreaks of SARS-CoV-2 in high schools. A simulation study in classrooms of four European countries. medRxiv. 2020.

[32] Keeling MJ, Tildesley MJ, Atkins BD, Penman B, Southall E, Guyver-Fletcher G, et al. The impact of school reopening on the spread of COVID-19 in England. medRxiv. 2020:2020.06.04.20121434. Available from: http://medrxiv.org/content/early/2020/09/16/2020.06.04.20121434.abstract.

[33] Munday JD, Jarvis CI, Gimma A, Wong KL, van Zandvoort K, Funk S, et al. Estimating the impact of reopening schools on the reproduction number of SARS-CoV-2 in England, using weekly contact survey data. medRxiv. 2021.

[34] Brooks-Pollock E, Read JM, McLean AR, Keeling MJ, Danon L. Mapping social distancing measures to the reproduction number for COVID-19. Philosophical Transactions of the Royal Society B. 2021;376(1829):20200276.

[35] UK Government: Prime Minister’s Office. Educational setting status; 2020. Available from: https://form.education.gov.uk/service/educational-setting-status.

[36] UK Government. Guidance: COVID-19 testing data: methodology note; 2021. Available from: https://www.gov.uk/government/publications/coronavirus-covid-19-testing-data-methodology/covid-19-testing-data-methodology-note.

[37] Lenormand M, Jabot F, Deffuant G. Adaptive approximate Bayesian computation for complex models. Computational Statistics. 2013;28(6):2777–2796.

[38] Office for National Statistics.Dataset: Estimates of the population for the UK, England and Wales, Scotland and Northern Ireland; 2021. Available from: https://www.ons.gov.uk/peoplepopulationandcommunity/populationandmigration/populationestimates/datasets/populationestimatesforukenglandandwalesscotlandandnorthernireland.

[39] Office for National Statistics. COVID-19 Schools Infection Survey Round 2, England: December 2020; 2021. Available from: https://www.ons.gov.uk/peoplepopulationandcommunity/healthandsocialcare/conditionsanddiseases/bulletins/covid19schoolsinfectionsurveyround2england/december2020.

[40] UK Government: Teaching Blog. How schools are managing bubbles effectively; 2021. Available from: https://teaching.blog.gov.uk/2020/09/28/how-schools-are-managing-bubbles-effectively/.

[41] Hart WS, Maini PK, Thompson RN. High infectiousness immediately before COVID-19 symptom onset highlights the importance of continued contact tracing. Elife. 2021;10:e65534.

[42] Ferretti L, Ledda A, Wymant C, Zhao L, Ledda V, AbelerDorner L, et al. The timing of COVID-19 transmission. medRxiv. 2020:2020.09.04.20188516. Available from: http://medrxiv.org/content/early/2020/09/16/2020.09.04.20188516.abstract.

[43] Lauer SA, Grantz KH, Bi Q, Jones FK, Zheng Q, Meredith HR, et al. The Incubation Period of Coronavirus Disease 2019 (COVID-19) From Publicly Reported Confirmed Cases: Estimation and Application. Annals of Internal Medicine. 2020 3.

[44] Davies NG, Klepac P, Liu Y, Prem K, Jit M, CMMID COVID-19 working group, et al. Age-dependent effects in the transmission and control of COVID-19 epidemics. Nat Med. 2020 aug;26(8):1205–1211. Available from: http://www.nature.com/articles/s41591-020-0962-9.

[45] McEvoy D, McAloon CG, Collins AB, Hunt K, Butler F, Byrne AW, et al. The relative infectiousness of asymptomatic SARS-CoV-2 infected persons compared with symptomatic individuals: A rapid scoping review. medRxiv. 2020:2020.07.30.20165084. Available from: http://medrxiv.org/content/early/2020/08/01/2020.07.30.20165084.abstract.

[46] Buitrago-Garcia D, Egli-Gany D, Counotte MJ, Hossmann S, Imeri H, Ipekci AM, et al. Occurrence and transmission potential of asymptomatic and presymptomatic SARS-CoV-2 infections: A living systematic review and meta-analysis. PLOS Med. 2020 sep;17(9):e1003346.

[47] Office for National Statistics. Coronavirus (COVID-19) Infection Survey pilot: England, Wales and Northern Ireland, 25 September 2020; 2020. Available from: https://www.ons.gov.uk/peoplepopulationandcommunity/healthandsocialcare/conditionsanddiseases/bulletins/coronaviruscovid19infectionsurveypilot/englandwalesandnorthernireland25september2020.

[48] Report written by: Andrew Rambaut, Nick Loman, Oliver Pybus, Wendy Barclay, Jeff Barrett, Alesandro Carabelli, Tom Connor, Tom Peacock, David L Robertson, Erik Volz, on behalf of COVID-19 Genomics Consortium UK (CoG-UK). Preliminary genomic characterisation of an emergent SARS-CoV-2 lineage in the UK defined by a novel set of spike mutations; 2020. Available from: https://virological.org/t/preliminary-genomic-characterisation-of-an-emergent-sars-cov-2-lineage-in-the-uk-defined-by-a-novel-set-of-spike-mutations/563.

[49] Riley S, Walters CE, Wang H, Eales O, Haw D, Ainslie KE, et al. REACT-1 round 12 report: resurgence of SARS-CoV-2 infections in England associated with increased frequency of the Delta variant. medRxiv. 2021.

[50] Public Health England. Stay at home: guidance for households with possible or confirmed coronavirus (COVID-19) infection.; 2021. Available from: https://www.gov.uk/government/publications/covid-19-stay-at-home-guidance/stay-at-home-guidance-for-households-with-possible-coronavirus-covid-19-infection.

[51] Hellewell J, Russell TW, Beale R, Kelly G, Houlihan C, Nastouli E, et al. Estimating the effectiveness of routine asymptomatic PCR testing at different frequencies for the detection of SARS-CoV-2 infections. BMC Med. 2021 dec;19:106. Available from: https://bmcmedicine.biomedcentral.com/articles/10.1186/s12916-021-01982-x.

[52] Office for National Statistics. Coronavirus (COVID-19) Infection Survey: antibody data for the UK, January 2021; 2021. Available from: https://www.ons.gov.uk/peoplepopulationandcommunity/healthandsocialcare/conditionsanddiseases/articles/coronaviruscovid19infectionsinthecommunityinengland/antibodydatafortheukjanuary2021.

[53] Department of Health and Social Care. NHS test and trace: how it works.; 2021. Available from: https://www.gov.uk/guidance/nhs-test-and-trace-how-it-works.

[54] Munday JD, Sherratt K, Meakin S, Endo A, Pearson CAB, Hellewell J, et al. Implications of the school-household network structure on SARS-CoV-2 transmission under school reopening strategies in England. Nat Commun. 2021 ec;12:1942. Available from: http://www.nature.com/articles/s41467-021-22213-0.

[55] Brooks S, Gelman A, Jones G, Meng XL, editors. Handbook of Markov Chain Monte Carlo. Chapman and Hall/CRC; 2011. Available from: https://www.taylorfrancis.com/books/9781420079425.

[56] Skittrall JP, Fortune MD, Jalal H, Zhang H, Enoch DA, Brown NM, et al. Diagnostic tool or screening programme? Asymptomatic testing for SARS-CoV-2 needs clear goals and protocols. The Lancet Regional Health-Europe. 2021;1.

[57] He X, Lau EHY, Wu P, Deng X, Wang J, Hao X, et al. Temporal dynamics in viral shedding and transmissibility of COVID-19. Nat Med. 2020 may;26(5):672–675. Available from: http://www.nature.com/articles/s41591-020-0869-5.

[58] Tupper P, Colijn C. COVID-19 in schools: Mitigating classroom clusters in the context of variable transmission. PLOS Computational Biology. 2021. Available from: https://doi.org/10.1371/journal.pcbi.1009120.

[59] Public Health England. SARS-CoV-2 variants of concern and variants under investigation in England. Technical briefing 14: 3 June 2021; 2021. Available from: https://www.ons.gov.uk/peoplepopulationandcommunity/healthandsocialcare/conditionsanddiseases/bulletins/coronaviruscovid19infectionsurveyantibodyandvaccinationdatafortheuk/latest.

[60] Stein-Zamir C, Abramson N, Shoob H, Libal E, Bitan M, Cardash T, et al. A large COVID-19 outbreak in a high school 10 days after schools’ reopening, Israel, May 2020. Eurosurveillance. 2020;25(29):2001352.

[61] Lessler J, Grabowski MK, Grantz KH, Badillo-Goicoechea E, Metcalf CJE, Lupton-Smith C, et al. Household COVID-19 risk and in-person schooling. medRxiv. 2021:2021.02.27.21252597. Available from: http://medrxiv.org/content/early/2021/03/01/2021.02.27.21252597.abstract.

[62] Salathé M, Kazandjieva M, Lee JW, Levis P, Feldman MW, Jones JH. A high-resolution human contact network for infectious disease transmission. Proceedings of the National Academy of Sciences. 2010;107(51):22020–22025.

[63] Eames KT, Tilston NL, Edmunds WJ. The impact of school holidays on the social mixing patterns of school children. Epidemics. 2011;3(2):103–108.

[64] Conlan AJ, Eames KT, Gage JA, von Kirchbach JC, Ross JV, Saenz RA, et al. Measuring social networks in British primary schools through scientific engagement. Proceedings of the Royal Society B: Biological Sciences. 2011;278(1711):1467–1475.

[65] Jarvis CI, Van Zandvoort K, Gimma A, Prem K, CMMID COVID-19 working group, Klepac P, et al. Quantifying the impact of physical distance measures on the transmission of COVID-19 in the UK. BMC Med. 2020 ec;18:124. Available from: https://bmcmedicine.biomedcentral.com/articles/10.1186/s12916-020-01597-8.

[66] Funk S, Munblit D, Flasche S. LFD mass testing in English schools: additional evidence of high test specificity. CMMID Repository. 2021. Available from: https://cmmid.github.io/topics/covid19/mass-testing-schools.html.

